# Genetic aetiologies for childhood speech disorder: novel pathways co-expressed during brain development

**DOI:** 10.1101/2022.05.15.22274630

**Authors:** Antony Kaspi, Michael S. Hildebrand, Victoria E. Jackson, Ruth Braden, Olivia van Reyk, Tegan Howell, Simone Debono, Mariana Lauretta, Lottie Morison, Matthew Coleman, Richard Webster, David Coman, Himanshu Goel, Mathew Wallis, Gabriel Dabscheck, Lilian Downie, Emma K. Baker, Bronwyn Parry-Fielder, Kirrie Ballard, Eva Harrold, Shaun Ziegenfusz, Mark F. Bennett, Erandee Robertson, Longfei Wang, Amber Boys, Simon E. Fisher, David J. Amor, Ingrid E. Scheffer, Melanie Bahlo, Angela T. Morgan

**Author notes:** These authors contributed equally to the work (Melanie Bahlo, Angela T. Morgan).

## Abstract

Childhood apraxia of speech (CAS), the prototypic severe childhood speech disorder, is characterized by motor programming and planning deficits. Genetic factors make substantive contributions to CAS aetiology, with a monogenic pathogenic variant identified in a third of cases, implicating around 20 single genes to date. Here we ascertained 70 unrelated probands with a clinical diagnosis of CAS and performed trio genome sequencing. Our bioinformatic analysis examined single nucleotide, indel, copy number, structural and short tandem repeat variants. We prioritised appropriate variants arising *de novo* or inherited that were expected to be damaging based on *in silico* predictions. We identified high confidence variants in 18/70 (26%) probands, almost doubling the current number of candidate genes for CAS. Three of the 18 variants affected *SETBP1*, *SETD1A* and *DDX3X*, thus confirming their roles in CAS, while the remaining 15 occurred in genes not previously associated with this disorder. Fifteen variants arose *de novo* and three were inherited. We provide further novel insights into the biology of child speech disorder, highlighting the roles of chromatin organization and gene regulation in CAS, and confirm that genes involved in CAS are co-expressed during brain development. Our findings confirm a diagnostic yield comparable to, or even higher, than other neurodevelopmental disorders with substantial *de novo* variant burden. Data also support the increasingly recognised overlaps between genes conferring risk for a range of neurodevelopmental disorders. Understanding the aetiological basis of CAS is critical to end the diagnostic odyssey and ensure affected individuals are poised for precision medicine trials.

## Introduction

Childhood apraxia of speech (CAS) is a rare neurodevelopmental disorder, occurring in 0.1% of the population ^1^. CAS stems from deficits in speech planning and programming, affecting a child’s ability to sequence sounds and syllables accurately and with correct prosody, resulting in highly unintelligible speech ^1, 2^. The first evidence implicating a specific gene in aetiology of CAS was provided in 2001, via a family study revealing that pathogenic variants in *FOXP2* were responsible for the speech disorder ^3^. For almost two decades, *FOXP2* was the only gene associated with CAS, in the absence of intellectual disability. Technological advances and reduced costs for DNA analyses have recently enabled efficient genome sequencing and bioinformatic follow-up, paving the way for high throughput discovery of genes involved in CAS. In particular, two independent cohort studies have performed genome-wide sequencing on 52 individuals with CAS ^4, 5^.

In the first cohort, aetiologic variants were identified in eight of 19 probands ascertained for CAS, yielding a genetic diagnostic rate of 42% with pathogenic variants revealed in: *CHD3*, *SETD1A*, *WDR5*, *KAT6A*, *SETBP1*, *ZFHX4*, *TNRC6B* and *MKL2* ^4^. In the second cohort, comprising 33 probands with CAS, nine additional genes were implicated: *CDK13, EBF3, GNAO1, GNB1, DDX3X, MEIS2, POGZ, UPF2* and *ZNF142*. One individual also had a contiguous gene deletion, yielding a genetic diagnostic rate of 33% (11/33) across this second cohort ^5^. In these studies, there was no evidence of recurrent point mutations and no genes which appeared to carry a higher burden of mutations, except for *SETBP1;* for which two individuals were found to carry variants across the two cohorts ^6^.

Taken together, the two cohort studies provided novel insights into the neurobiology of childhood speech disorders. First, the discovery of 17 new genes involved in CAS aetiology, with a combined diagnostic yield of 37%, revealing for the first time, that many children do have a single gene diagnosis explaining their speech condition. Second, many of the highly penetrant variants implicated shared pathways broadly involved in transcriptional regulation (e.g. *POGZ, SETBP1, SETD1A, KAT6A*), suggesting a key role for transcriptional dysregulation in aberrant speech development^4, 5^. Other molecular pathways of significance were also were also revealed with high confidence and likely novel pathogenic variants, such as in *GNA01* and *GNB1,* both part of G-protein signaling pathways ^5^. Third, the studies demonstrated that pathogenic variants more commonly arise *de novo* rather than being inherited, and that speech disorders are genetically heterogeneous ^5^, as is the case for other neurodevelopmental disorders ^7–9^. Fourth, the candidate genes newly implicated in CAS were frequently associated with other neurodevelopmental disorders, such as epilepsy (e.g. *GNAO1, GNB1, SETD1A*) and/or intellectual disability (e.g. *CDK13*, *CHD3*, *DDX3X, POGZ, SETBP1*) ^4, 5^. These novel insights into CAS aetiology, including genetic heterogeneity, demonstrate the need to study additional, larger cohorts to reveal further causative genes, increase the genetic diagnostic yield, and further unravel molecular pathways underlying severe childhood speech disorder. A much deeper knowledge of the molecular bases of severe speech conditions such as CAS is essential to move the field toward precision therapies.

Here, we aimed to identify molecular bases in a large cohort of probands ascertained for a primary diagnosis of CAS. Each proband underwent comprehensive phenotypic analysis and genome sequencing to identify pathogenic variants of major effect. We also analysed the molecular co-expression of all genes associated with CAS, and the overlap of genes associated with CAS and other neurodevelopmental disorders.

## Methods

### Ethical consent

The study was approved by the Human Research Ethics Committee of The Royal Children’s Hospital, Melbourne, Australia (#37353). Written informed consent was obtained from parents or legal guardians to take part in the study and have the results of this study published.

### Participants and phenotyping

Probands under age 18 years were ascertained with a clinical diagnosis of CAS and where parents and clinicians reported the current primary clinical concern as poor speech development^5^. Probands with moderate to severe intellectual disability were excluded. Participants were recruited via medical and speech pathology clinicians or direct parent referral. Medical and developmental history and secondary neurodevelopmental outcomes were recorded with validation via relevant professional reports (e.g. paediatrician, multi-disciplinary assessment team for ASD diagnosis, physiotherapist, occupational therapist, academic outcomes) (Tables 1, 2; Supplementary Table 1).

**Table 1:** Medical and neurodevelopmental features of individuals with CAS and pathogenic/likely pathogenic variants

| Family | Age, y;m | Sex | Core speech phenotype | Gene | Gross motor delay | Fine motor delay | Vision impaired<sup>#</sup> | Hearing loss<sup>#</sup> | MRI findings<sup>#</sup> | Seizures<sup>#</sup> | Other NDD<sup>#</sup> | Dysmorphic features<sup>#</sup> | Other medical<sup>#</sup> |
| --- | --- | --- | --- | --- | --- | --- | --- | --- | --- | --- | --- | --- | --- |
| 1 | 15-17 | F | CAS | *ARHGEF9* | Y | Y |  |  |  |  |  |  |  |
| 2 | 6-8 | F | Dysarthria | *DDX3X* | Y | Y |  |  |  |  |  |  |  |
| 3 | 6-8 | F | CAS | *KDM5C* | N | N |  |  |  |  |  |  |  |
| 4 | 6-8 | M | CAS | *PHF21A* | Y | Y |  |  |  |  |  |  |  |
| 5 | 3-5 | M | Inconsistent phonological delay and disorder | *BRPF1* | Y | N |  |  |  |  |  |  |  |
| 6 | 6-8 | F | Phonological disorder, articulation disorder | *PURA* | Y | Y |  |  |  |  |  |  |  |
| 7 | 3-5 | M | CAS | *ZBTB18* | Y | Y |  |  |  |  |  |  |  |
| 8 | 3-5 | F | CAS | *HNRNPK* | Y | Y |  |  |  |  |  |  |  |
| 9 | 6-8 | M | CAS, phonological disorder | *SETD1A* | Y | Y |  |  |  |  |  |  |  |
| 10 | 9-11 | M | CAS,<br>phonological<br>error patterns | *SETD1B* | Y | Y |  |  |  |  |  |  |  |
| 11 | 3-5 | M | CAS,<br>phonological<br>error patterns | *RBFOX3* | Y | Y |  |  |  |  |  |  |  |
| 12 | 3-5 | M | CAS,<br>phonological<br>error patterns | *TAOK2* | Y | Y |  |  |  |  |  |  |  |
| 13 | 0-1 | M | CAS,<br>minimally<br>verbal | *SPAST* | Y | N |  |  |  |  |  |  |  |
| 14 | 3-5 | M | CAS,<br>minimally<br>verbal | *SHANK3* | Y | Y |  |  |  |  |  |  |  |
| 15 | 6-8 | M | CAS,<br>phonological<br>delay | *DIP2C* | Y | Y |  |  |  |  |  |  |  |
| 16 | 3-5 | M | CAS | *SETBP1* | Y | Y |  |  |  |  |  |  |  |
| 17 | 6-8 | F | CAS,<br>Dysarthria | *ERF* | Y | Y |  |  |  |  |  |  |  |
| 18 | 3-5 | M | CAS | *TRIP12* | N | N |  |  |  |  |  |  |  |

NDD, Neurodevelopmental disorder; ASD, Autism spectrum disorder; CAS, Childhood apraxia of speech; DCD, Developmental coordination disorder; F, Female; ID, Intellectual disability; M, Male; N, Feature not present; SPD, Sensory processing disorder; Y, Feature present, #These details have been removed to avoid the identification of patients, ~Ages have been denoted in 2-year bands to avoid the identification of participants

**Table 2:** Linguistic phenotype and educational setting of individuals with CAS and pathogenic/likely pathogenic variants

| Family | Oral motor impairment | History of feeding issues | Language: receptive* | Language: expressive* | Reading deficits | Spelling deficits | Speech pathology | IQ<sup>#</sup> | Education setting |
| --- | --- | --- | --- | --- | --- | --- | --- | --- | --- |
| 1 | Y | Y | Severe | Severe | Y | Y | Y |  | Specialist |
| 2 | Y | Y | Severe | Severe | Y | Y | Y |  | Mainstream (with classroom support) |
| 3 | Y | Y | Moderate | Severe | Y | Y | Y |  | Mainstream |
| 4 | Y | N | Severe | Severe | Y | Y | Y |  | Mainstream |
| 5 | Y | Y | Average | Moderate | TY | TY | Y |  | Mainstream |
| 6 | Y | Y | Severe | NA | Y | Y | Y |  | Mainstream (with classroom support) |
| 7 | Y | Y | Severe | Severe | TY | TY | Y |  | Mainstream |
| 8 | Y | Y | NS | NS | TY | TY | Y |  | Not yet at school |
| 9 | Y | N | Moderate | Moderate | Y | Y | Y |  | Mainstream |
| 10 | Y | N | Average | Average | Y | Y | Y |  | School for children with specific speech and language impairments |
| 11 | Y | N | Mild | Moderate | TY | TY | Y |  | Mainstream |
| 12 | Y | N | Severe | Mild | TY | TY | Y |  | Specialist |
| 13 | Y | N | Average | Severe | TY | TY | Y |  | Mainstream |
| 14 | Y | Y | Severe | Severe | TY | TY | Y |  | Mainstream |
| 15 | Y | N | Average | Moderate | Y | Y | Y |  | Mainstream |
| 16 | Y | N | Severe | Severe | TY | TY | Y |  | Mainstream |
| 17 | Y | Y | Moderate | Severe | TY | TY | Y |  | Mainstream |
| 18 | Y | N | Average | Average | TY | TY | Y |  | Mainstream |

FSIQ, Full scale IQ; N, No; NA, Not assessed; VCI: Verbal Comprehension Index; VSI: Visual Spatial Comprehension; GMDS: Griffiths
Mental Developmental Scales; WNV: Weschler non-verbal scale of ability; KBIT: Kaufman Brief Intelligence Test; PRI, Perceptual reasoning
index; TY, not applicable for literacy testing as pre-school age (< 5 years old); Y, feature present.
\*Language severity rated according to CELF-5<sup>14</sup> as follows: 86-114 average, 78-85 mild, 71-77 moderate, <70 severe.
^ Assessment not previously indicated by the family or treating physician at time of study.

A diagnosis of CAS was based on meeting the three American Speech-Language-Hearing Association consensus criteria for CAS: 1) inconsistent errors on consonants and vowels in repeated productions of syllables or words; 2) lengthened and disrupted coarticulatory transitions between sounds and syllables, and 3) inappropriate prosody ^1^. Criteria were operationally defined and rated ^10^ from phonetic transcriptions of standardised single word speech sub-tests (phonology and inconsistency) ^11^ and a 5-minute conversational speech sample ^5^. Dysarthria was diagnosed in the presence of oral tone or coordination disturbance and dysarthric features identified during conversation using the Mayo Clinic Dysarthria rating scale ^12, 13^. Language and cognition were also assessed with standardised tools ^14–17^.

### Genetic testing

Genomic DNA was extracted from whole blood or saliva using a Qiagen (Valencia, CA) QIAamp DNA Maxi kit or a prepIT L2P kit (DNA Genotek Inc., Ontario, Canada), respectively. Probands underwent chromosomal microarray testing on Illumina (SanDiego, CA) platforms, with the reportable effective resolution of arrays being 200 Kb. Results were analyzed with Karyostudio software version 1.3 or 1.4 (Illumina), using genome reference sequence NCBI36/hg18 (v1.3, pre-2013) or GRCh37/hg19 (v1.4, 2013 onwards).

Genome sequencing was conducted on 204 individuals from 70 families comprising 71 probands (two probands were monozygotic twins hence for the genetic analysis and results we report on 70 probands), 127 parents and 6 other relatives. Illumina TruSeq DNA Nano or NovaSeq PE150 PCR free library preparation was completed prior to sequencing on the Illumina NovaSeq 6000 to average 30-fold depth with ∼100Gb data generated per sample at the Australian Genome Research Facility or Novogene (HK) Company Limited. Sanger sequencing or droplet digital PCR (ddPCR) were used to segregate variants in additional family members who had not undergone microarray or genome sequencing.

### Variant analysis

Variant discovery was performed using trio or parent–child pair (where one parent was unavailable for testing) designs (Fig. 1). Exceptions to this were two singletons, and four larger families. 150 bp sequence pair-end reads were mapped to the hg19 reference genome using the Burrow-Wheeler Aligner (BWA-MEM, bwa v0.7.17) ^18^. Read sorting and indexing was undertaken using SAMtools (v1.9) and Genome Analysis Toolkit (GATK, v4.1.4.1) was used to mark duplicates. Base quality score recalibration was performed, and variants were called using HaplotypeCaller, with individuals called separately, as implemented by GATK. Sequencing quality control was performed using fastQC.

**Fig. 1.**
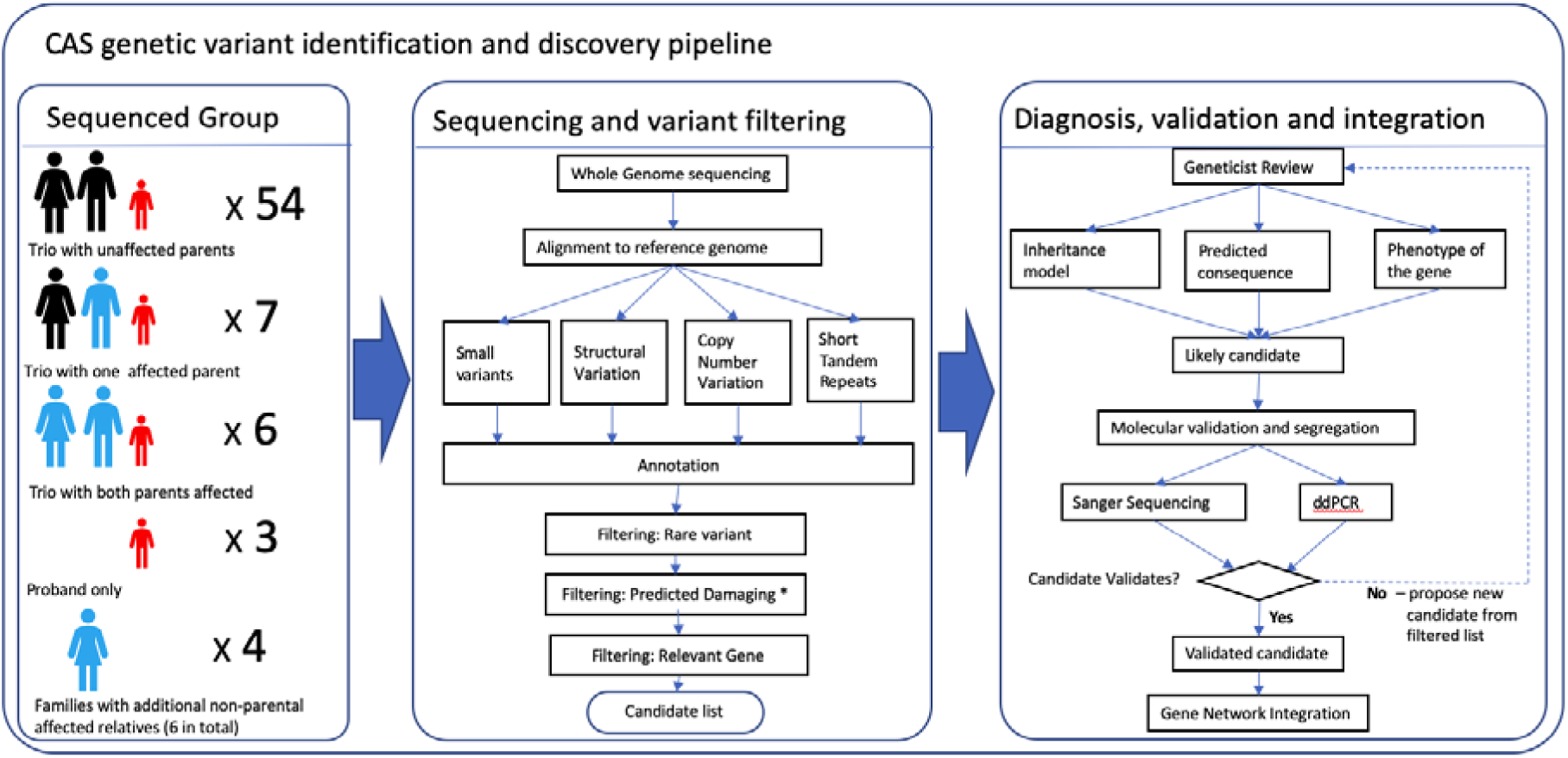
Genetic variant identification and variant filtering pipeline for individuals with CAS. Workflow covers recruitment of patients (CAS in red, affected relative in blue, unaffected in black), DNA sequencing, analysis and filtering of genomic data, identification of potential causative variants, geneticist review, molecular validation, segregation and integration of all findings. *Only the damaging effects of small variants are predicted bioinformatically.

Genotype calling and quality filtering were performed separately in multiple genome sequencing batches. Joint calling was performed by merging per-sample gvcf files and applying GATK’s GenotypeGVCFs tool. Variants with excess heterozygosity (Z score >4.5) were removed, then variant quality score recalibration was carried out for single nucleotide variants (SNVs) and indels separately, with a truth sensitivity filter of 99.7 to flag variants for exclusion. Filtering of low quality SNV calls excluded those flagged by low threshold or any of the following filters: low quality by depth (QD <2); evidence of strand bias (Fisher strand [FS] >60 or strand odds ratio [SOR] >3); and evidence of significant differences between alternate and reference alleles for read mapping qualities (rank sum < −12.6) or position bias (ReadPosRankSum < −8). Indels filtering was performed in a similar manner to missense variant filtering, with exceptions being to exclude variants with FS >200; SOR >10; or ReadPosRankSum < −20. Finally, familial relationships were confirmed using peddy ^19^. Filtering and other scripted analysis was conducted using R version 3.5.2.

Analysis was restricted to variants: (1) not present in gnomAD or with gnomAD allele count ≤ 2, (2) not present in unaffected family members from our cohort, and (3) potentially *de novo*, or consistent with an appropriate inheritance model matching the phenotypic pedigree (e.g. dominant, recessive). Compound heterozygous models were considered for variants present in gnomAD with a mean allele frequency <0.05%. Only variants with read depth >10 and genotype quality >20 in the proband and their sequenced family members were considered. Identified variants were annotated with the variant effect predictor (VEP v93.3) algorithm, using the assembly version GRCh37.p13 and categorized based on the following series of annotations.

### Genome-wide analysis of LoF and predicted damaging missense variants

We analyzed the genome sequencing data for loss of function (LoF) and predicted damaging missense variants genome-wide. Predicted LoF candidates were defined by using VEP annotations that were required to meet three criteria: (1) annotated as frameshift, stop or start lost, stop gained, splice acceptor or donor variant, (2) in a gene predicted intolerant to LoF variation (ExACpLI ≥0.9 or LoFtool <0.1), and (3) at least one of the following: (a) CADD Phred score ≥20 predicted damaging, or (b) predicted to affect splicing (AdaBoost score ≥0.6 or random forest score ≥0.6 using the dbscSNV VEP plugin). For frameshift variants, the variant was only required to be in a LoF intolerant gene.

Predicted damaging missense variants had to meet at least three criteria: (1) PolyPhen-2 prediction as “probably” or “possibly damaging”, (2) SIFT prediction as “deleterious” or “deleterious low confidence”, (3) a CADD Phred score ≥20 predicted damaging, or (4) a missense tolerance ratio significantly different from 1 (false discovery rate <0.05).

### Criteria for identification and reporting of candidate variants

We applied a two-stage approach for shortlisting candidate variants, from our identified LoF and damaging missense variants: (1) we selected variants located within genes of interest, a gene list collated based on several relevant criteria, informed by previous CAS studies^4, 5^, and described below. Pathogenicity of these variants was assessed using the American College of Medical Genetics (ACMG) guidelines, and via review by a clinical geneticist; (2) where no candidate variant was identified, we then applied a genome-wide, wholly agnostic to gene, search for candidate variants, to be followed up with ACMG and clinical geneticist review. The size of the cohort and the inability to perform statistical analyses to implicate novel genes, such as via burden analysis, necessitated the usage of these constraints.

We report candidate variants as follows:

1. High-confidence variants: LoF and predicted damaging missing variants, that were classified with the American College of Medical Genetics (ACMG) guidelines as pathogenic (class 5) or likely pathogenic (class 4) ^20^, and where the phenotype associated with the gene was consistent with that of the proband.
2. Low confidence variants: LoF and predicted damaging missing variants, that were either classified as of uncertain significance (3) according to ACMG guidelines, or classified likely pathogenic (class 4), but where the gene was not consistent with the proband’s phenotype, or otherwise lacked evidence for pathogenicity.

All reported variants were inspected with the Integrative Genome Viewer (v 2.7).

### Collated list of genes of interest

The list of genes of interest, used in shortlisting candidate variants (n=2145 genes, Supplementary Table 2), was collated from the following sources: genes from recent CAS cohort studies ^4, 5^(n=19 genes), or previously confirmed single genes implicated in CAS such as *FOXP2* or *GRIN2A* ^3, 21^ (n=81). Additionally, high-confidence genes known to harbour pathogenic variants in intellectual disability (n=1399), epilepsy (n=611), autism spectrum disorder (ASD, n=131) and cleft palate (n=156), recognised by Victorian Clinical Genetics Services, were extracted from PanelApp using an application programming interface (https://panelapp.agha.umccr.org/) ^22^. High-confidence ASD-related genes from the Simons Foundation Autism Research Initiative database were also included ^23^ (n=419). Finally, brain-expressed genes associated with primate-human accelerated evolution were included; this set comprised of 415 genes overlapping human accelerated regions (HARs) that are also significantly over-expressed in brain, compared to other tissues ^24^, and 45 genes overlapping with HARs, that were find to be exclusively expressed in human brain cells, and not in other primates ^25^.This final set of genes were included, as HARs have previously been implicated in ASD and cognitive development, and thus may be involved in the evolutionary development of speech.

### Copy number and structural variants

Manta (regions up to 5Mb) ^26^ (v 1.6.0) and qDNAseq (bin size 10kb with CNVs up to 5Mb) ^27^ (v 1.18.0) were used to detect CNVs and other structural variants. Manta detects structural variants based on abnormal alignment of read pairs. qDNAseq detects structural variants based on read depth. Variants occurring in more than two families were filtered out to avoid false positives due to technical artefact. SVAnnot (v 2.5) was used to annotate the variants, filtering by gnomAD SV abundance with SVs with frequency >0.05% excluded. Candidate structural variants were identified using the same approach as for SNVs, with pathogenicity assessed via ACMG guidelines and clinical review.

### Variant validation

High-confidence variants were validated using Sanger sequencing or ddPCR. For Sanger sequencing, gene variants were amplified using gene specific primers (oligonucleotide sequences available on request) designed to the reference human gene transcripts (NCBI Gene). Amplification reactions were cycled using a standard protocol on a Veriti Thermal Cycler (Applied Biosystems, Carlsbad, CA) at 60°C annealing temperature for 1 minute. Bidirectional sequencing of all exons and flanking regions was completed with a BigDye v3.1 Terminator Cycle Sequencing Kit (Applied Biosystems). Sequencing products were resolved using a 3730xl DNA Analyzer (Applied Biosystems). All sequencing chromatograms were compared to the published cDNA sequence; nucleotide changes were detected using Codon Code Aligner (CodonCode Corporation, Dedham, MA). For ddPCR, probes and primers were designed in-house and synthesized by Integrated DNA Technologies (Coraville, IA) and assays were performed ^28, 29^, using a Bio-Rad QX200 Droplet Digital PCR System (Hercules, CA) and QuantaSoft software v1.7.4.0917.

### Analysis of novel sources of genetic contributions to CAS

Three forms of genetic analysis for CAS that have not been previously applied were undertaken: (1) short tandem repeat (STR) analysis of both known and novel pathogenic repeats (Supplementary Table 3); (2) examination of common variants implicated in ASD, cleft palate, and word reading ability, and their relevance to CAS, via associations with polygenic risk scores (PRS) and (3) estimation of mitochondrial gene abundance (see Supplementary methods).

### Brain gene co-expression and gene set enrichment analysis

Gene co-expression analyses were undertaken for our high-confidence genes (n=18) and then extended to include 16 genes previously implicated in CAS ^4, 5^, totalling 34 genes. Analyses were conducted using a Monte Carlo sampling approach ^4, 5^ with data from the BrainSpan Atlas of the Developing Human Brain ^30^ (Supplementary Table 4). Co-expression analyses were also performed to prioritize genes of uncertain significance for CAS (see Supplementary methods).

Gene set enrichment analyses were undertaken using gene sets in Gene Ontology molecular function, cellular component, and biological processes databases as well as Reactome pathway databases ^31^. g:Profiler was used to test for gene set enrichment ^32^ with a Bonferroni-corrected p-value threshold of 0.05 to determine pathways enriched for genes implicated in CAS.

### Data availability

Data can be made available by contacting the corresponding author.

## Results

### Phenotypic data

The 71 probands (53 males, 18 females, 1 monozygotic twin pair) from the 70 families had an average age of 5 years 7 months (range 2 years 2 months to 16 years 8 months). Phenotypic information for the cohort is shown in Tables 1, 2; Supplementary Tables 1a, 1b. Pathogenic variants were confirmed in 18 probands (12 males, 6 females; average age of 5 years 7 months) (Tables 1, 2) as described further in the following section. The phenotypes of the probands with (n = 18) versus without (n = 52) variants are presented in Fig. 2.

**Fig. 2:**
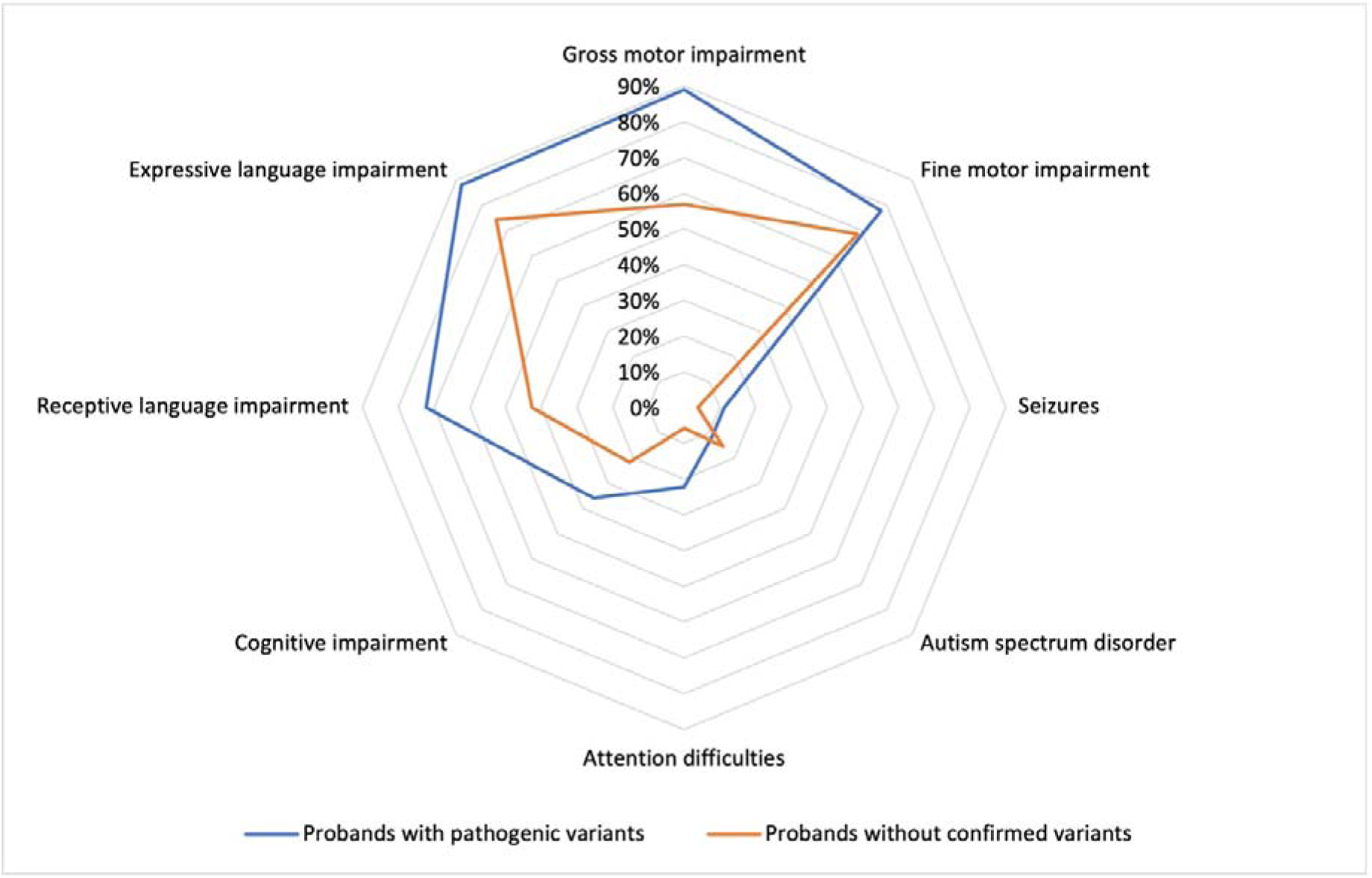
Phenotypic overlap in childhood apraxia of speech (CAS) cohort. Phenotypic features of CAS cohort with (n=18) and without (n=52) pathogenic variants. Data based on children with formal assessments by health professionals (i.e., cognition, language, motor, formal ASD diagnoses). Data from Tables 1, 2; Supplementary Tables 1a, 1b.

All probands were ascertained based on a clinical diagnosis of CAS. Following our speech assessment protocol, 15 of the 18 probands with pathogenic variants had CAS in isolation (n = 7) or co-occurring with other speech disorders (n = 8) (Table 1). Three probands (IDs 2, 5, and 6; Table 1) had other severe speech disorder presentations (dysarthria, n = 1; phonological and articulation disorder, n = 1; inconsistent phonological disorder, n = 1). Expressive language disorder was implicated in 15/17 probands (mild, n = 1; moderate, n = 4; severe or unable to be scored due to severity, n = 10; Fig. 2). One proband was minimally verbal and unable to complete a valid expressive language assessment. Receptive language disorder was noted in 13/18 individuals (mild, n = 1; moderate, n = 3; severe, n = 9). Of those old enough to read and write (> 5 years, n = 8), 8 had reading impairment, and 8 had spelling impairment. Two probands with pathogenic variants (2/18, 11%; IDs 10, 18) had CAS accompanied by fine motor and related linguistic deficits, but without other neurodevelopmental disorder diagnoses.

Of the 18 children identified to carry high confidence variants, 14 had formal cognitive assessment with profiles ranging from an average full-scale IQ (FSIQ) (n = 3), to borderline FSIQ results (n = 6), to mild intellectual disability (n = 5). For 3 children, FSIQ could not be calculated due to significant performance variation across verbal and nonverbal subscales, which is a common experience for children with severe speech production deficits. The remainder (n = 4) did not have IQ testing because concerns with learning or cognition had not been raised or pursued by the family or treating physician and children were attending mainstream childcare or school settings (IDs 13-16; IDs 13, 14, 16 were < 4 years of age when few children receive formal cognitive testing). Other features included neurodevelopmental diagnoses or features secondary to CAS including mild ASD (n = 2), difficulties with attention (n = 4), and anxiety and mood-related symptoms (n = 1). Dysmorphic features such as epicanthic folds and pointed chin (Table 1), rated by a clinical geneticist with 24 years of clinical experience, were present in just over half of the probands with high confidence variants (11/18). Gross motor (n = 16) and fine motor delays (n = 14) were common and associated with a slower trajectory in learning to ride a bicycle, balance appropriately, draw, write, and cut compared to typical peers. Two of the 18 children with pathogenic variants (IDs 1, 12) had a history of seizures.

### Single nucleotide and indel variants

A high confidence variant was identified in 18/70 (26%) of probands (Table 3a, Fig. 3). These included three frameshift, two splice acceptor, six nonsense, and six missense variants, as well as one multiple exon duplication, and they were found in 18 different genes (*ARHGEF9*, *BRPF1, DDX3X, DIP2C, ERF, HRNPNK, KDM5C, PHF21A, PURA, RBFOX3, SETBP1, SETD1A, SETD1B, SHANK3, SPAST, TAOK2, TRIP12, ZBTB18)*. All high-confidence variants were *de novo* except in *PURA, ERF* and *RBFOX3,* which were inherited (Table 3a, Fig. 3). Many of these genes that we newly implicate in CAS, as well as genes previously described in earlier sequenced CAS cohorts^4, 5^ are also associated with other neurodevelopmental disorders (Fig. 4a and Fig. 4b, Supplementary Table 5).

**Fig. 3.**
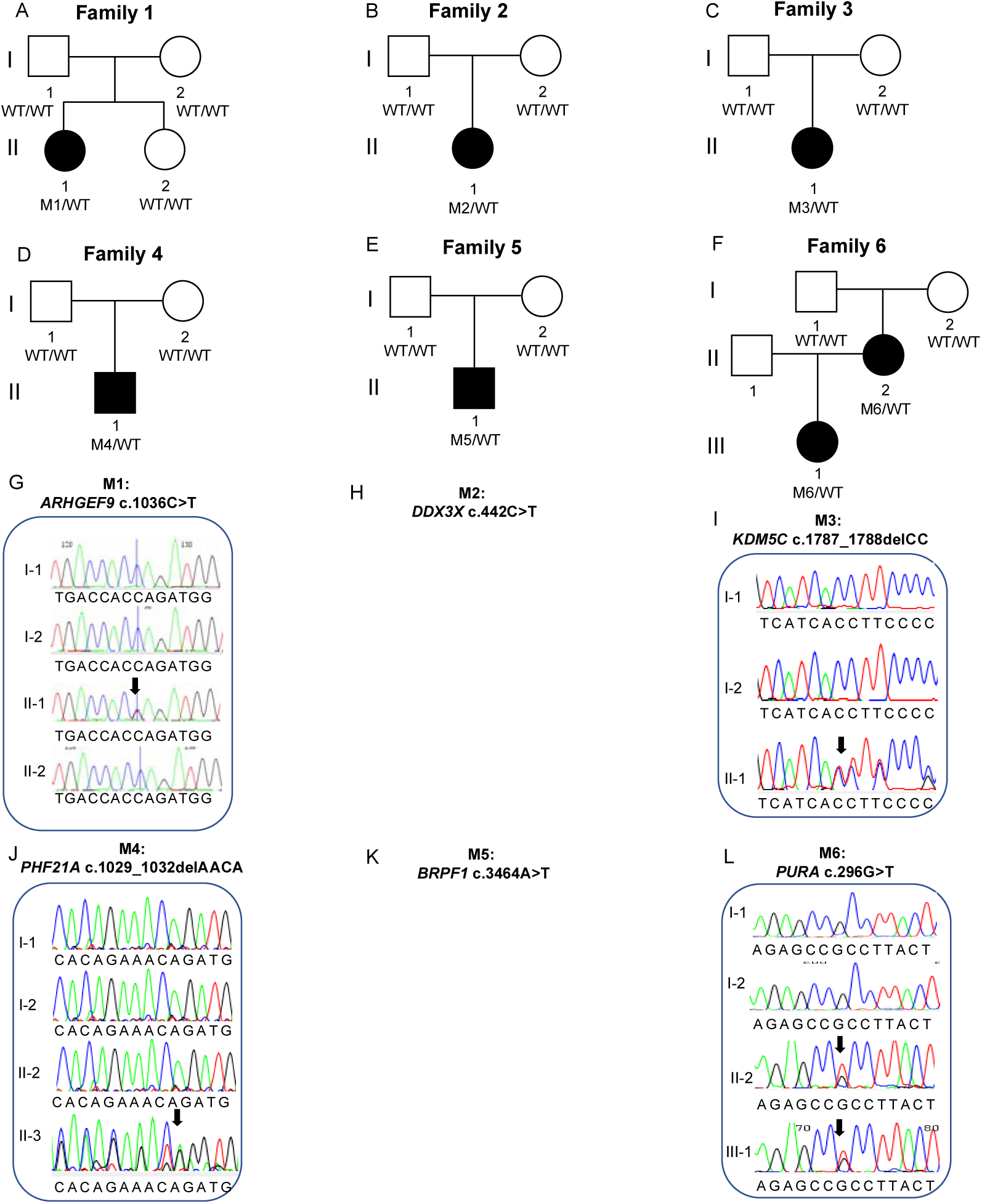
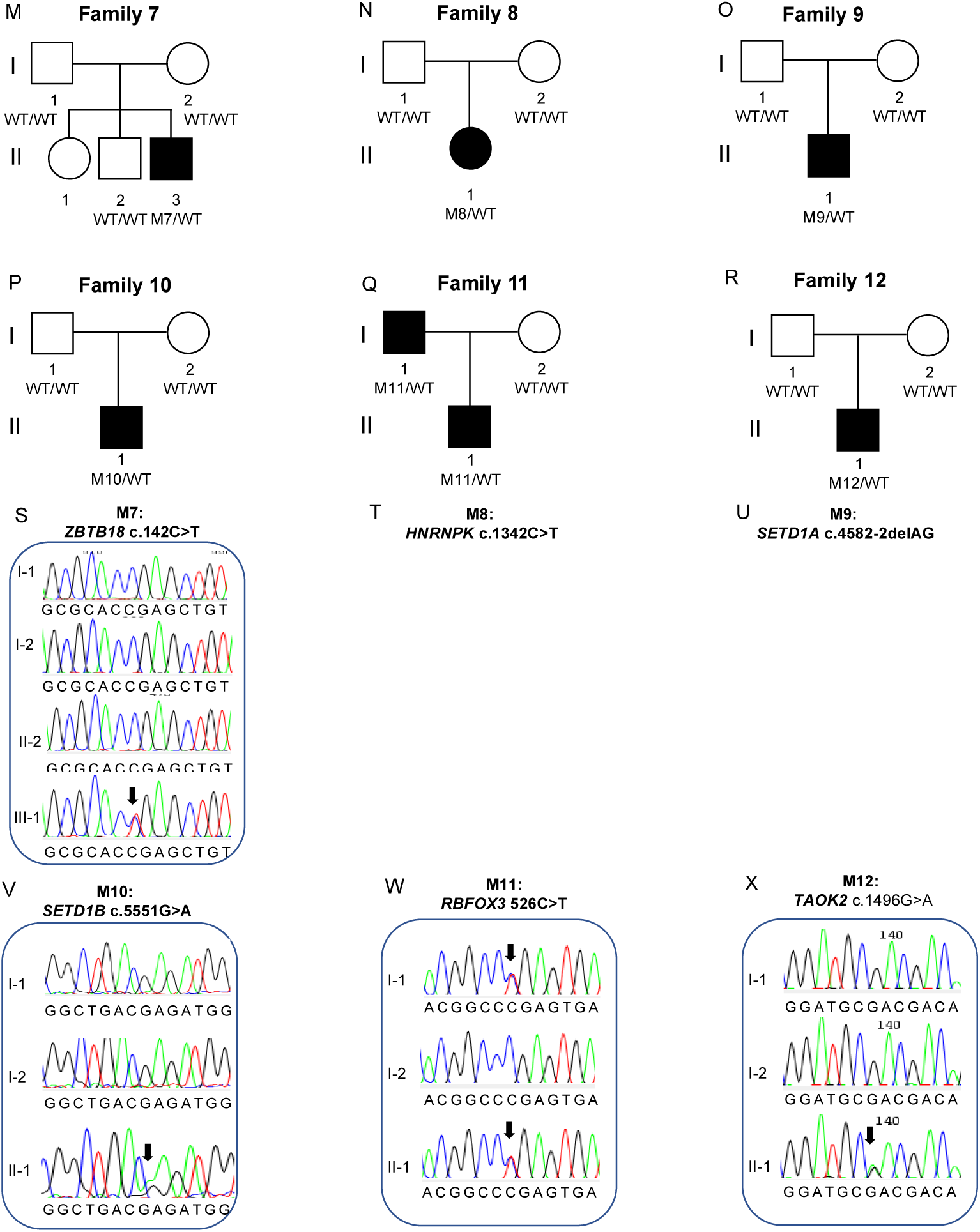
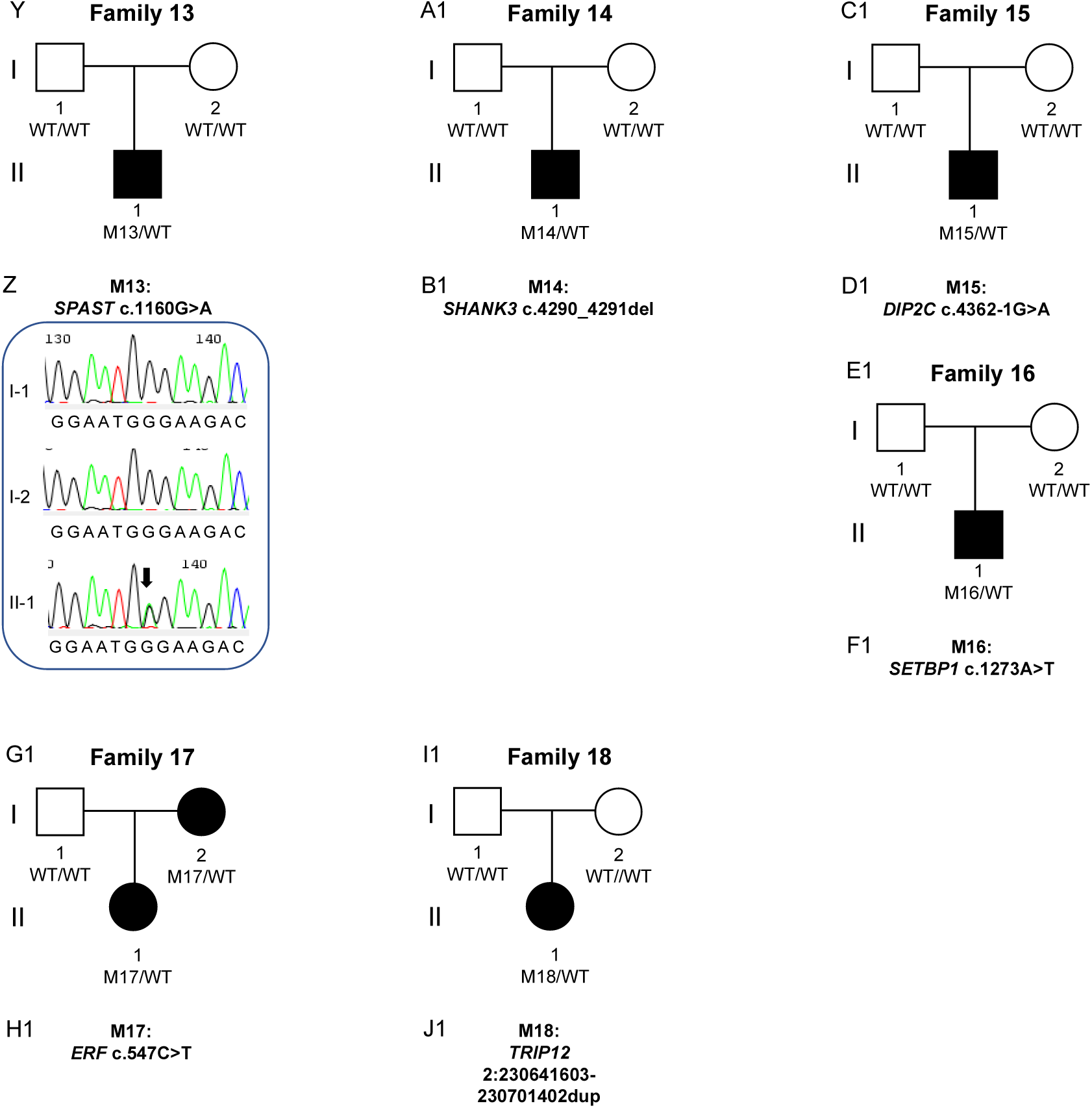
Families with High Confidence Variants analysed by genome sequencing. Families analysed by Genome Sequencing. Pedigrees (A-F, M-R, Y, A1, C1, E1, G1, I1) from 18 families with 18 different high confidence variants. Sequence chromatograms (G, I, J, L, S, T, V, W, X, Z) showing *de novo* or inherited variants. Sanger sequencing was not performed for the variants in eight of the families (B, E, O, A1, C1, E1, G1, I1).

**Fig. 4:**
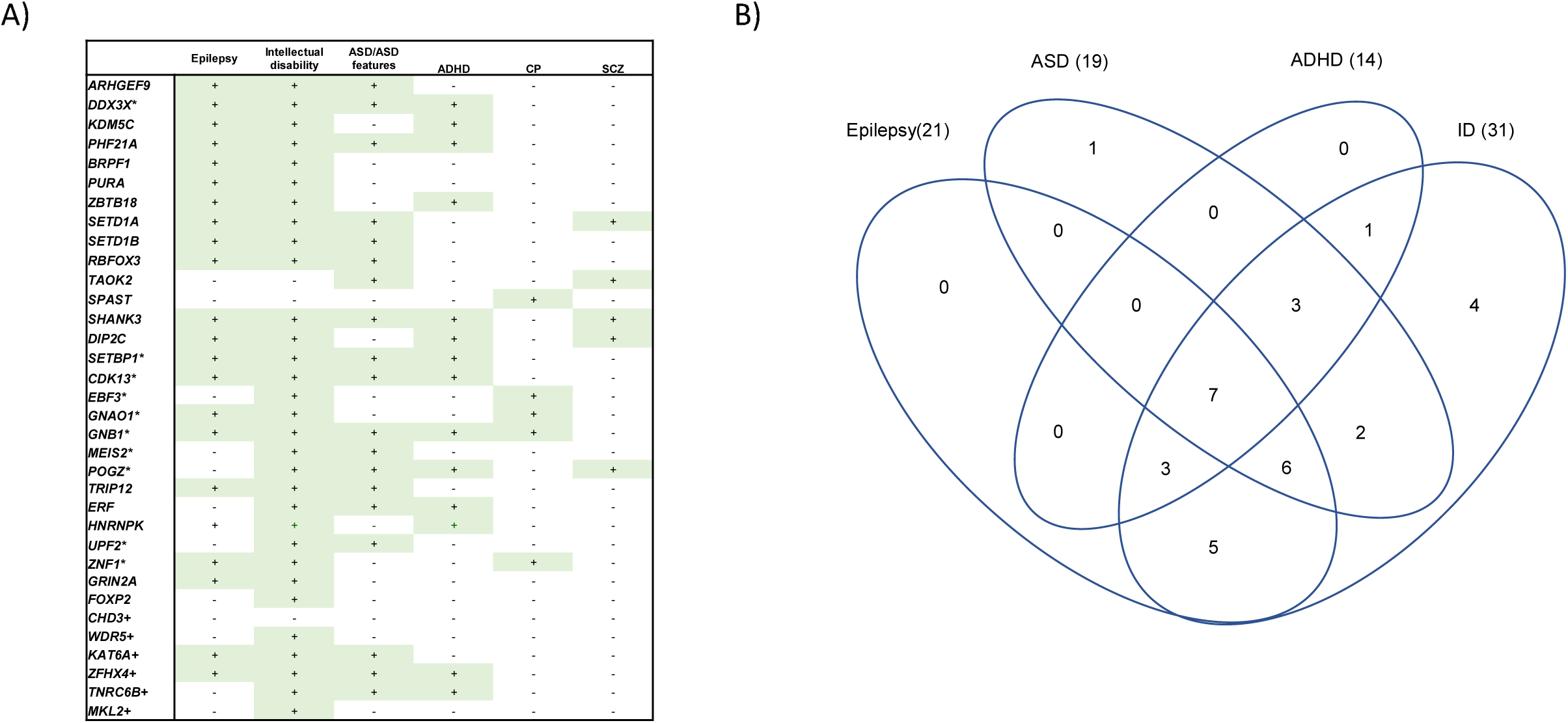
Previously identified neurodevelopmental conditions in candidate genes for CAS. (A) Candidate Genes for CAS identified in this study and Hildebrand et. al. (*) also have been shown to cause other neurodevelopmental disorder traits. (B) Venn diagram showing the overlap of these genes and multiple neurodevelopmental disorder traits.

**Table 3.** a: High-confidence gene variants in individuals with CAS

| Family | Sex | Chr:Pos | Gene | OMIM | DNA Variant | Protein Change | Effect | In Silico Predictions<sup>s</sup> | gnomAD Count<sup>~</sup> | Inheritance | ACMG score | Reference<sup>*</sup> |
| --- | --- | --- | --- | --- | --- | --- | --- | --- | --- | --- | --- | --- |
| 1 | F | X:62885786 | ARHGEF9 | 616056 | c.1036C>T | p.Q346* | Nonsense | gnomADpLI = 0.98;<br>LoFtool = 0.0652;<br>CADD= 38 | 0 | De novo | PVS1, PS2, PM2, PP3<br>Class 5<br>Pathogenic |  |
| 2 | F | X:41201905 | DDX3X | 300958 | c.442C>T | p.Q148* | Nonsense | gnomADpLI = 1<br>LoFtool = 0.0555;<br>CADD= 43 | 0 | De novo | PVS1, PM2, PP3<br>Class 5<br>Pathogenic |  |
| 3 | F | X:53231113 | KDM5C | 300534 | c.1787_1788delC<br>C | p.T596IfsTer25 | Frameshift | gnomADpLI = 1;<br>LoFtool = 0.0636; | 0 | De novo | PVS1, PS2, PM2, PP3<br>Class 5<br>Pathogenic |  |
| 4 | M | 11:45975138 | PHF21A | 608325 | c.1029_1032delA<br>ACA | p.T344RfsTer28 | Frameshift | gnomADpLI = 1;<br>LoFtool = 0.08; | 0 | De novo | PVS1, PS2, PM2, PP3<br>Class 5<br>Pathogenic |  |
| 5 | M | 3:9788123 | BRPF1 | 617333 | c.3464A>T | p.D1155V | Missense | SIFT = Del(0);<br>PolyPhen = Dam (0.99);<br>CADD= 33 | 0 | De novo | PS2, PM2, PP3, BP1<br>Class 4 Likely<br>Pathogenic |  |
| 6 | F | 5:139494062 | PURA | 616158 | c.296G>T | p.R99L | Missense | SIFT = Del(0);<br>PolyPhen = Dam (0.984);<br>CADD= 28.7 | 0 | Inherited from affected mother | PM2, PP1, PP2, PP3<br>Class 4 Likely<br>Pathogenic |  |
| 7 | M | 1:244217218 | ZBTB18 | 612337 | c.142C>T | p.R48* | Nonsense | gnomADpLI = 1<br>LoFtool = 0.18;<br>CADD= 36 | 0 | De novo | PVS1, PS2, PM2, PP3, PP5<br>Class 5<br>Pathogenic | <sup>34</sup> |
| 8 | F | 9: 86585096 | *HNRNPK* | 616580 | c.1342C>T | p.Q448* | Nonsense | gnomADpLI = 1<br>CADD= 43 | 0 | *De novo* | PS2, PM2, PP3.<br>Class 5<br>Pathogenic |  |
| 9 | M | 16:30992057 | *SETD1A* | 619056 | c.4582-2delAG | NA | Splice<br>Acceptor Site | gnomADpLI = 1<br>LoFtool = 0.00654;<br>CADD= 24.6 | 2 | *De novo* | PVS1, PS2,<br>PM2, PP3<br>Class 5<br>Pathogenic | <sup>33</sup> |
| 10 | M | 12:22265722 | *SETD1B* | 619000 | c.5551G>A | p.E1851K | Missense | SIFT = Del(0);<br>PolyPhen = Dam (1);<br>CADD= 33 | 0 | *De novo* | PS2, PM1, PM2,<br>PP2, PP3<br>Class 5<br>Pathogenic |  |
| 11 | M | 17:77097708 | *RBFOX3* | 616999 | c.526C>T | p.R176* | Nonsense | gnomADpLI = 1<br>LoFtool = 0.17;<br>CADD= 46 | 0 | Inherited<br>from<br>affected<br>father | PVS1, PM2, PP1<br>Class 5<br>Pathogenic |  |
| 12 | M | 16:29996607 | *TAOK2* | 613199 | c.1496G>A | p.R499Q | Missense | SIFT = Del(0.01);<br>PolyPhen = Dam (0.992);<br>CADD= 32 | 0 | *De novo* | PS2, PM2, PP3,<br>BP1<br>Class 4 Likely<br>Pathogenic |  |
| 13 | M | 2:32352078 | *SPAST* | 182601 | c.1160G>A | p.G387E | Missense | SIFT = Del(0);<br>PolyPhen = Dam (1);<br>CADD= 32 | 0 | *De novo* | PS2, PM1, PM2,<br>PP2, PP3<br>Class 5<br>Pathogenic |  |
| 14 | M | 22:51160325 | *SHANK3* | 606232 | c.4290_4291del | p.V1432Gfs*4 | Frameshift | gnomADpLI =1<br>CADD= 35 | 0 | *De novo* | PVS1, PS2,<br>PM2, PP3, PP5<br>Class 5<br>Pathogenic | <sup>66</sup> |
| 15 | M | 10: 323518 | *DIP2C* | 611380 | c.4362-1G>A | NA | Splice<br>Acceptor Site | gnomADpLI =1<br>LoFtool =0.356;<br>CADD= 26.1 | 0 | *De novo* | PVS1, PS2,<br>PM2, PP3<br>Class 5<br>Pathogenic |  |
| 16 | M | 18: 42530578 | *SETBP1* | 611060 | c.1273A>T | p.K425* | Nonsense | gnomADpLI = 1<br>LoFtool = 0.<br>0.0297;<br>CADD= 38 | 0 | *De novo* | PVS1, PS2,<br>PM2, PP3<br>Class 5<br>Pathogenic |  |
| 17 | M | 19:42753717 | ERF | 617180 | c.547C>T | p.R183* | Nonsense | gnomADpLI = 0.99<br>LoFtool = 0.02;<br>CADD= 35 | 0 | Inherited from affected Mother | PVS1, PM2, PP1, PP5 Class 5 Pathogenic | <sup>36</sup> |
| 18 | F | 2:230641603-230701402dup | TRIP12 | 617752 | NA | NA | Exonic duplication | gnomADpLI = 1 | 0 | De novo | PVS1, PS2, PM2, PP3, PP5 Class 5 Pathogenic |  |

**Table 3.**
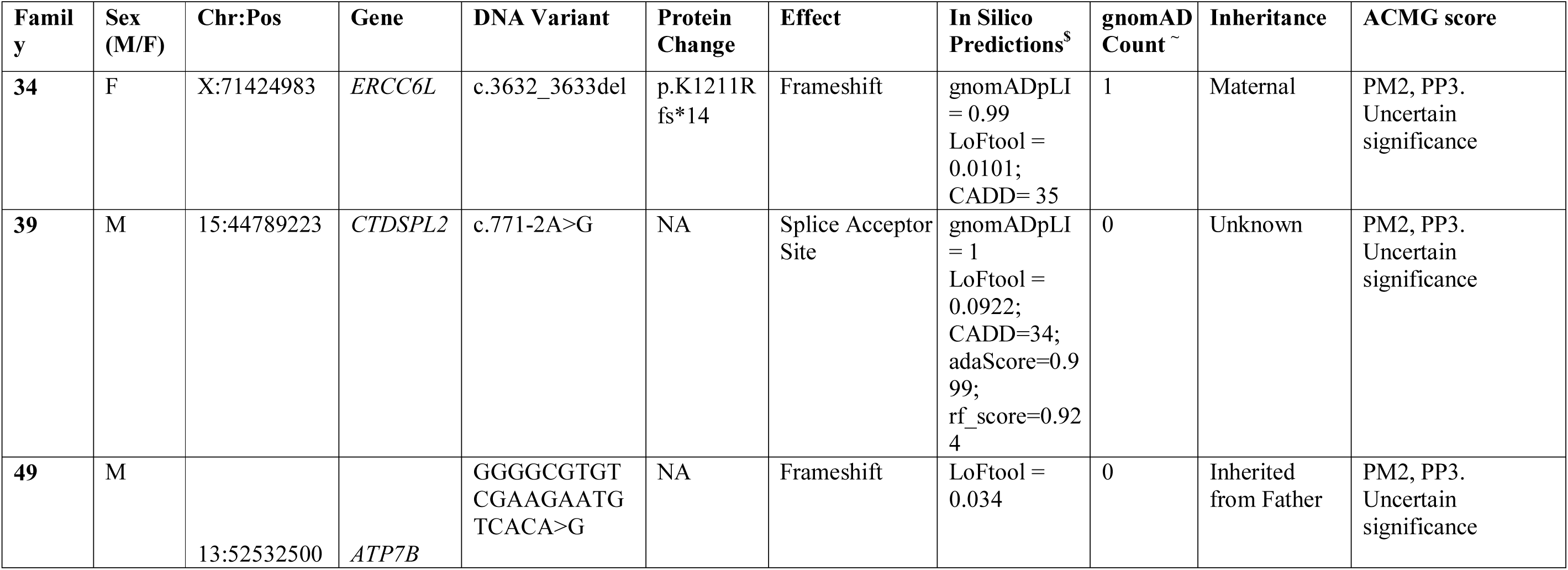
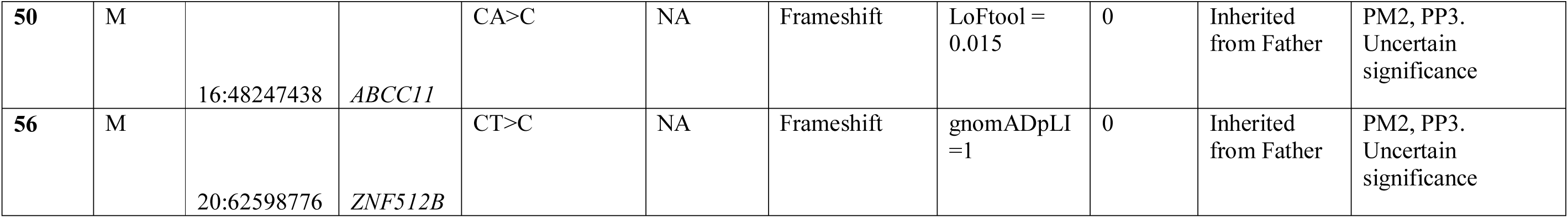
b: Low confidence, predicted LoF variants in individuals with CAS

**Table 3.**
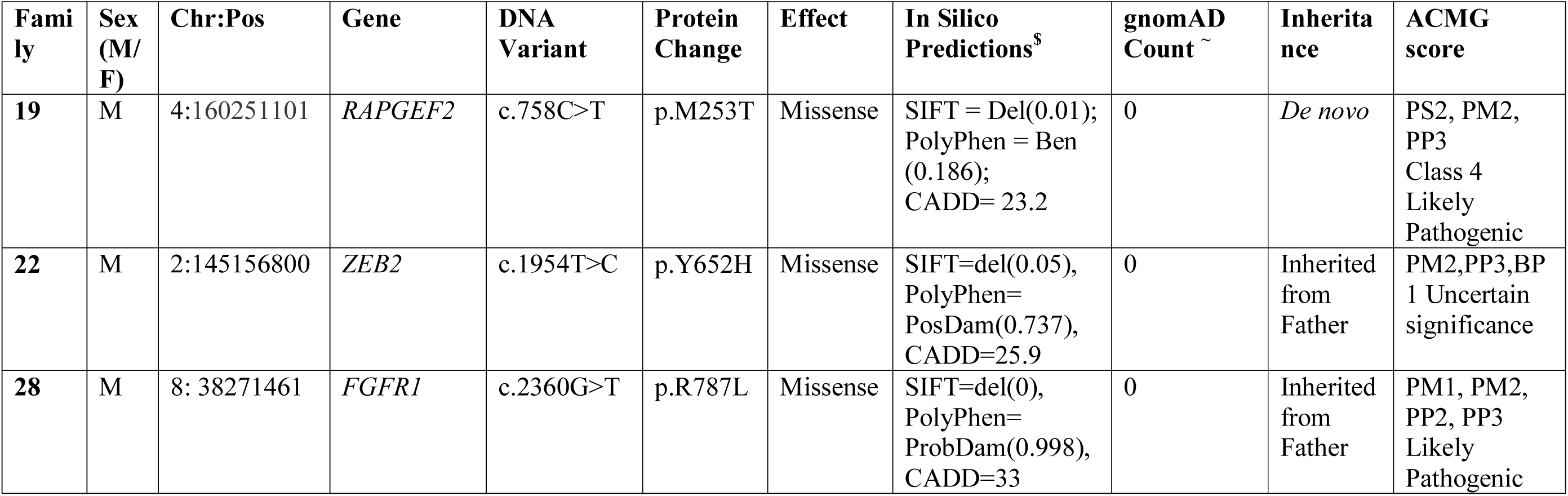
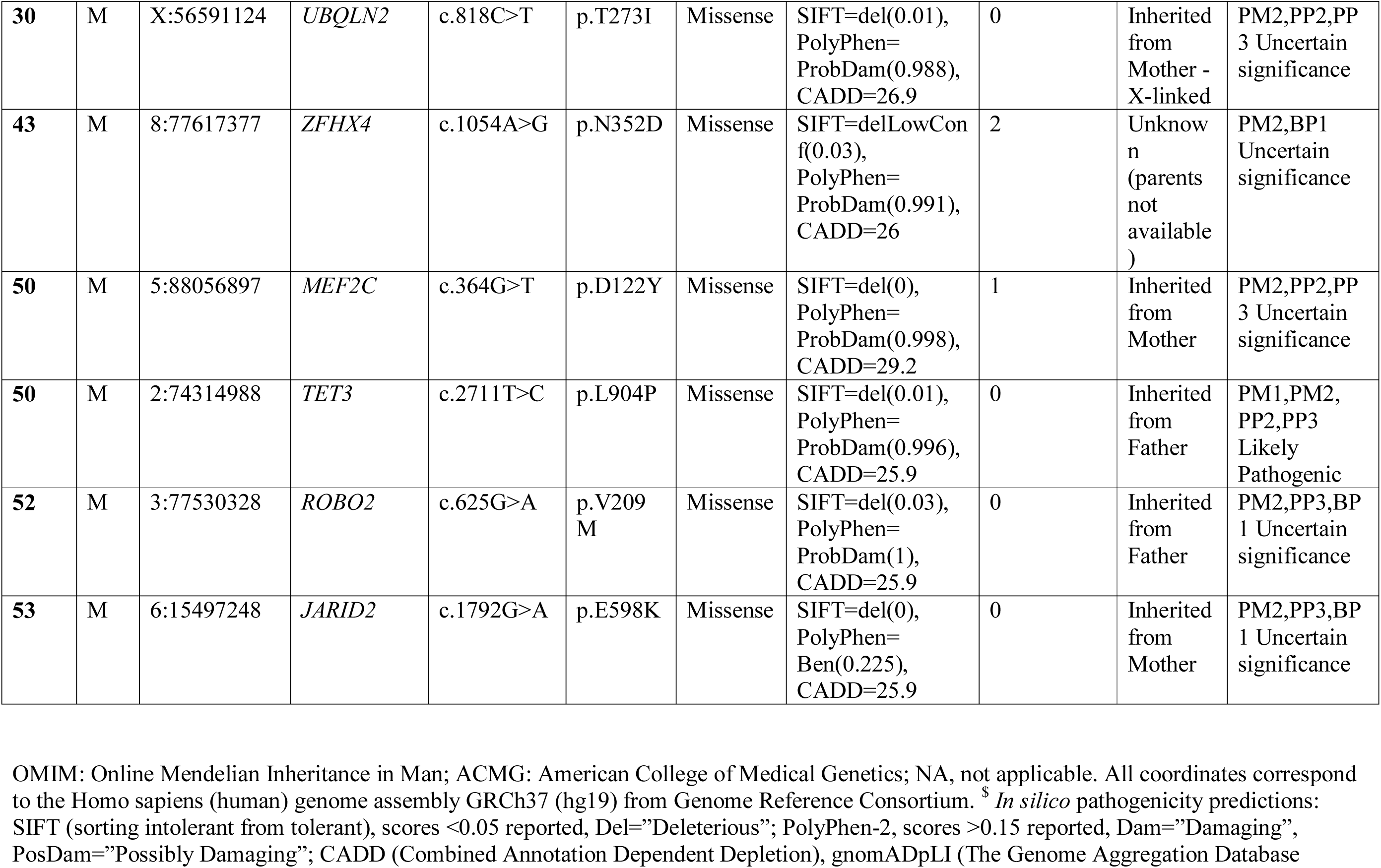
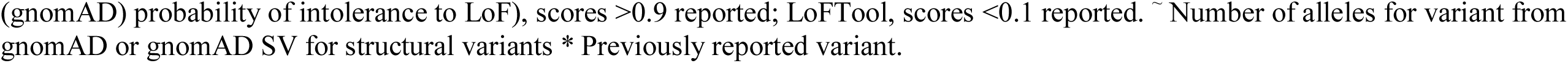
c: Low confidence, predicted damaging Missense variants in individuals with CAS

| **Fami**<br>**ly** | **Sex**<br>**(M/**<br>**F)** | **Chr:Pos** | **Gene** | **DNA**<br>**Variant** | **Protein**<br>**Change** | **Effect** | **In Silico**<br>**Predictions<sup>\$</sup>** | **gnomAD**<br>**Count ~** | **Inherita**<br>**nce** | **ACMG**<br>**score** |
| --- | --- | --- | --- | --- | --- | --- | --- | --- | --- | --- |
| **19** | M | 4:160251101 | *RAPGEF2* | c.758C>T | p.M253T | Missense | SIFT = Del(0.01);<br>PolyPhen = Ben<br>(0.186);<br>CADD= 23.2 | 0 | *De novo* | PS2, PM2,<br>PP3<br>Class 4<br>Likely<br>Pathogenic |
| **22** | M | 2:145156800 | *ZEB2* | c.1954T>C | p.Y652H | Missense | SIFT=del(0.05),<br>PolyPhen=<br>PosDam(0.737),<br>CADD=25.9 | 0 | Inherited<br>from<br>Father | PM2,PP3,BP<br>1 Uncertain<br>significance |
| **28** | M | 8: 38271461 | *FGFR1* | c.2360G>T | p.R787L | Missense | SIFT=del(0),<br>PolyPhen=<br>ProbDam(0.998),<br>CADD=33 | 0 | Inherited<br>from<br>Father | PM1, PM2,<br>PP2, PP3<br>Likely<br>Pathogenic |
| 30 | M | X:56591124 | *UBQLN2* | c.818C>T | p.T273I | Missense | SIFT=del(0.01), PolyPhen= ProbDam(0.988), CADD=26.9 | 0 | Inherited from Mother - X-linked | PM2,PP2,PP3 Uncertain significance |
| 43 | M | 8:77617377 | *ZFHX4* | c.1054A>G | p.N352D | Missense | SIFT=delLowConf(0.03), PolyPhen= ProbDam(0.991), CADD=26 | 2 | Unknown (parents not available) | PM2,BP1 Uncertain significance |
| 50 | M | 5:88056897 | *MEF2C* | c.364G>T | p.D122Y | Missense | SIFT=del(0), PolyPhen= ProbDam(0.998), CADD=29.2 | 1 | Inherited from Mother | PM2,PP2,PP3 Uncertain significance |
| 50 | M | 2:74314988 | *TET3* | c.2711T>C | p.L904P | Missense | SIFT=del(0.01), PolyPhen= ProbDam(0.996), CADD=25.9 | 0 | Inherited from Father | PM1,PM2,PP2,PP3 Likely Pathogenic |
| 52 | M | 3:77530328 | *ROBO2* | c.625G>A | p.V209M | Missense | SIFT=del(0.03), PolyPhen= ProbDam(1), CADD=25.9 | 0 | Inherited from Father | PM2,PP3,BP1 Uncertain significance |
| 53 | M | 6:15497248 | *JARID2* | c.1792G>A | p.E598K | Missense | SIFT=del(0), PolyPhen= Ben(0.225), CADD=25.9 | 0 | Inherited from Mother | PM2,PP3,BP1 Uncertain significance |

OMIM: Online Mendelian Inheritance in Man; ACMG: American College of Medical Genetics; NA, not applicable. All coordinates correspond to the Homo sapiens (human) genome assembly GRCh37 (hg19) from Genome Reference Consortium. <sup>\$</sup> *In silico* pathogenicity predictions: SIFT (sorting intolerant from tolerant), scores <0.05 reported, Del="Deleterious"; PolyPhen-2, scores >0.15 reported, Dam="Damaging", PosDam="Possibly Damaging"; CADD (Combined Annotation Dependent Depletion), gnomADpLI (The Genome Aggregation Database
(gnomAD) probability of intolerance to LoF), scores >0.9 reported; LoFTool, scores <0.1 reported. ~ Number of alleles for variant from gnomAD or gnomAD SV for structural variants \* Previously reported variant.

The 13 nonsense, splice-site, frameshift or exon duplication variants were all in genes intolerant to loss-of-function variation (*ARHGEF9*, *DDX3X, DIP2C, ERF, HNRNPK, KDM5C, PHF21A, RBFOX3, SETD1A, SETBP1, SHANK3, TRIP12, ZBTB18)*, according to gnomADpLI or LoFtool scores (Table 3a). The five missense variants were all predicted to be damaging by at least three in silico tools (SIFT, PolyPhen, CADD, MTR). Four of the 18 high confidence variants (in *ERF, SETD1A, SHANK3* and *ZBTB18)* were recurrent, with these variants having been reported previously in individuals with neurodevelopmental disorders ^33–37^. The remaining 14 high-confidence variants have not been previously reported: of these, ten were pathogenic and four were likely pathogenic ^20^.

In five probands, we identified low confidence, LoF variants in genes predicted to be intolerant to haploinsufficiency (Table 3b, *ABCC11, ATP7B, CTDSPL2, ERCC6L, ZNF512B*). These variants are all predicted to cause loss of function of the gene; however at present, none of these genes are established to cause CAS or other neurodevelopmental disorders and therefore are variants of unknown significance. Of note, a frameshift variant in *ATP7B* of uncertain significance, was also identified in our previous CAS cohort ^5^; in the present cohort, the identified variant (proband 49) is shared with their father; however, both parents of this proband are affected by CAS, so the variant does not fully segregate with CAS status. Thus, the relevance of *ATP7B* in CAS remains unclear.

In eight probands (8/70; 11.4%), we report nine rare (gnomAD allele count <=2) low confidence, predicted damaging missense variants (Table 3c). These are a selected subset of predicted damaging variants, located in genes that were of relevance due to known disease significance, or biological relevance, but were of uncertain pathogenicity, or the gene was not consistent with the proband’s phenotype. Two of the nine variants were in genes previously associated with speech and/or language disorders - *ROBO2*, and *ZFHX4*; however, one of these was inherited from an unaffected parent (*ROBO2*, Proband 52), and for the other, it was not possibly to determine whether the variant was de novo, as parental DNA was unavailable (*ZFHX4,* proband 43). The remaining seven variants were located in *CHD5*, *FGFR1*, *JARID2*, *MEF2C*, *RAPGEF2, TET3*, *UBQLN2* and *ZEB2*. For each of these, the variant was deemed to be of uncertain significance (ACMG), and/or the phenotype associated with the gene was not consistent with the proband’s phenotype. For all probands without a high confidence variant, the remaining predicted damaging missing variants, identified though our genome-wide search, are listed in Supplementary Table 6.

### Copy number and structural variation

There were no diagnostic findings on clinical microarray analysis. In one individual (ID18) a *de novo* pathogenic tandem 59,799bp duplication was identified in *TRIP12*, spanning exons 7 to 37 out of 42(NM_001348323.3). This duplication is predicted to disrupt the reading frame causing loss of function (Supplementary Fig. 1).

### Analysis of novel sources of genetic contributions to CAS

No repeat expansion of either known or novel repeats were identified in the CAS probands.

The polygenic risk score analysis did not identify any statistically significant findings, with the strongest trend being observed for ASD where probands were enriched for ASD risk, nearly achieving nominal significance (Two sample t-test, p=0.054). The cleft palate PRS also showed increased risk for CAS probands, but this was not significant (two sample t-test, p=0.226). Word reading was also decreased in CAS probands (two sample t-test, p=0.123).

Mitochondrial abundance analyses identified two CAS probands with high confidence, likely pathogenic variants in genes known to have mitochondrial function as outliers (*DDX3X* and *HNRNPK*) but overall mitochondrial abundance did not appear to be a biomarker for CAS overall (see Supplementary results, Supplementary Fig. 2A-C).

### Brain gene co-expression and gene set enrichment analyses

The median absolute correlation between our 18 high-confidence genes was |ρ| = 0.4194 (Fig. 5A). Thirty-two of the 153 pairwise correlations were among the top 5% most highly correlated gene pairs genome-wide |ρ| > 0.647, Fig. 5B), and there was evidence that this set of genes was more highly co-expressed than expected by chance (p = 0.0038). Gene set enrichment analyses of a subset of seven highly co-expressed genes (*BRPF1, DIP2C, KDM5C, PHF21A, SETBP1, SETD1A, SETD1B*) indicate they are involved in chromatin organization (GO:0006325; p = 1.238 x 10^-3^). (Supplementary Table 7)

**Fig. 5:**
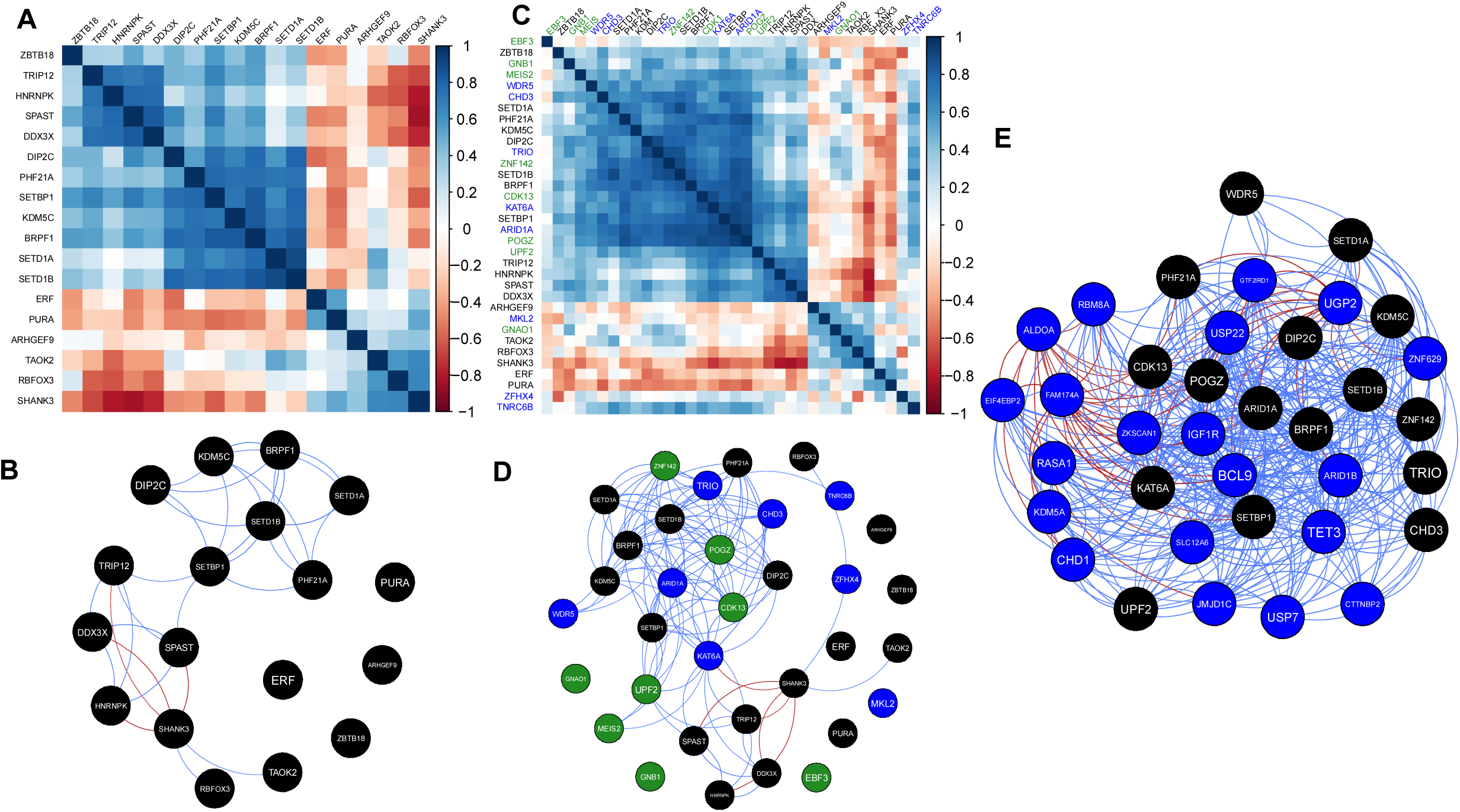
CAS candidate gene co-expression. (A) Gene co-expression matrix for the 18 high-confidence candidate genes with pairwise Spearman’s rank correlation coefficients between genes shown, based on 280 samples from 24 individuals (8 weeks post conception to 10 months after birth) from the BrainSpan resource. Genes ordered by hierarchical clustering, using the median linkage method. (B) Network of gene co-expression. Nodes represent genes; edges represent gene–pair correlations that exceed the threshold for the top 5% most highly correlated gene pairs genome-wide (|ρ| > 0.64) (blue – positive correlation, red – negative correlation). (C) Gene co-expression matrix for the 18 high- confidence novel candidate genes (black) as well as the genes from ^4^ (green) and ^5^ (blue). (D) Network of gene co-expression. Nodes represent genes; edges represent gene–pair correlations that exceed the threshold for the top 5% most highly correlated gene pairs genome-wide (|ρ| > 0.64) (blue – positive correlation, red – negative correlation). Black nodes - novel genes from this work, green nodes genes from ^4^and blue nodes from ^5^ (E) Network of gene co-expression. Nodes represent genes; edges represent gene–pair correlations that exceed the threshold for the top 5% most highly correlated gene pairs genome-wide (|ρ| > 0.64) (blue – positive correlation, red – negative correlation). Black nodes are a set of co-expressed genes including novel genes from this work as well as those from earlier works^4, 5^. Blue nodes - the top prioritized genes from cytogenic variants described in Table 4.

**Table 4.**
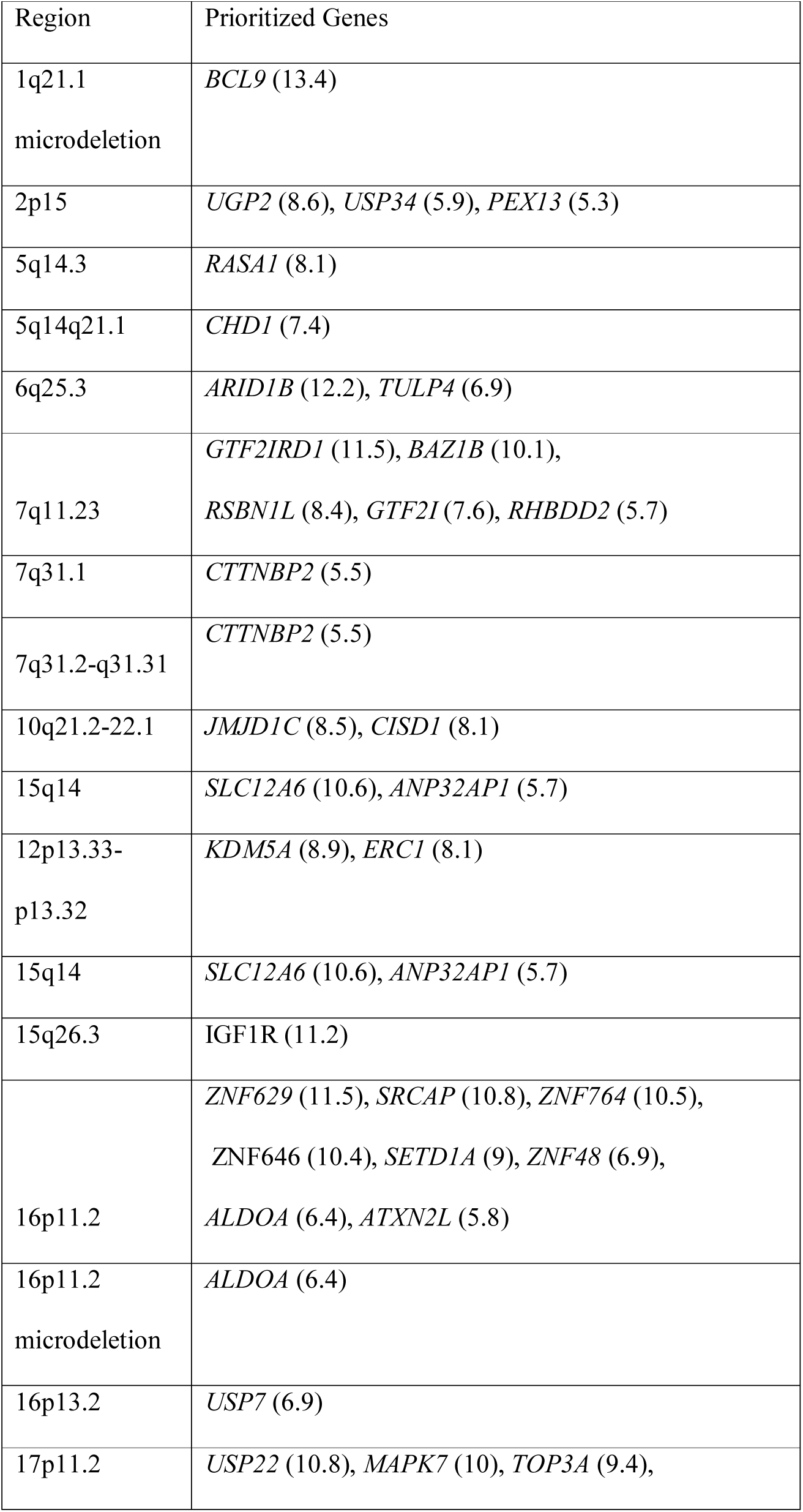
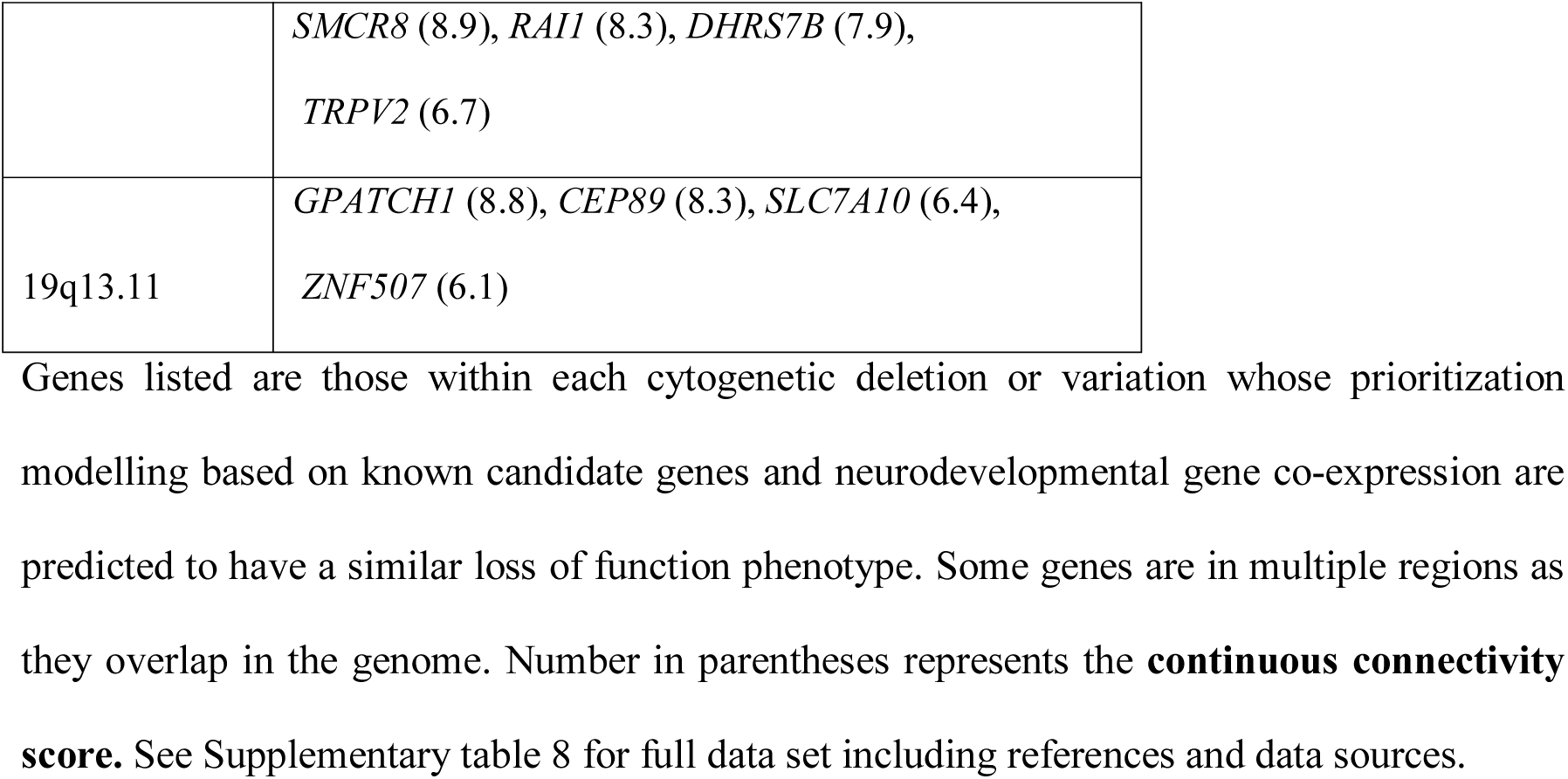
Genes prioritized in cytogenetic regions which have previously been identified with speech and language disorders

The median pairwise correlation of gene expression for the 34 genes (18 high-confidence genes from the current study and 16 genes previously implicated in CAS), was significantly higher than expected by chance (median |ρ| = 0.4095, p = < 2 x 10^-4^, Fig. 5D). Gene set enrichment analyses of the highly co-expressed cluster of 15 genes from the present study and past cohorts ^4, 5^ (Fig. 5C) further highlighted the significant over-representation of genes involved in chromatin organization (GO:0006325; Bonferroni-corrected p = 2.304 x 10^-6^) as well as transcriptional regulation (GO:0003676; Bonferroni-corrected p = 1.103 x 10^-4^, 25,396 sets tested, Supplementary Table 7).

Finally, our co-expression model was used to prioritize candidate genes, beyond our high confidence set. Firstly we re-examined the set of low-confidence variants identified in the present study (Table 3b and c), and variants of uncertain significance from our previous cohort.^5^ Amongst the low confidence findings in this paper, *TET3* was the only gene identified for proritization (FDR<0.1), while three genes identified in our previous cohort were prioritized (*BRWD3, MCMBP* and *ZKSCAN1*). All four prioritzed genes are associated with chromatin organization and/or DNA binding. Second, we sought to prioritize genes contained in each of 21 large copy number variant regions, identified through a literature search. All regions span multiple genes, and the associated phenotypes include speech disorder as an associated clinical feature (Supplementary Tables 8 and 9). Prioritization analysis identified at least one gene in each region (FDR <0.1), with more than one candidate for 18/21 regions (Table 4) (Fig. 4E). In several instances, the prioritized gene from our co-expression network had already been proposed as the likely causal gene (see Supplementary results).

## Discussion

Our findings almost double the current number of genes implicated in susceptibility to CAS and provide further novel insights into the biology of childhood speech disorder. We identified high confidence variants, thereby providing a clinical genetic diagnosis, for 18 children ascertained for CAS, revealing 15 genes that have not previously been associated with severe speech disorder (*ARHGEF9*, *BRPF1, DDX3X, DIP2C, ERF, HRNPNK, KDM5C, PHF21A, PURA, RBFOX3, SETBP1, SETD1A, SETD1B, SHANK3, SPAST, TAOK2, TRIP12, ZBTB18*). We identified a clinical genetic diagnosis in one-quarter of individuals tested; a diagnostic yield comparable to or even higher than other neurodevelopmental disorders with substantial *de novo variant burden*^38^. We provide independent confirmation with unrelated cases for three genes previously shown to have causative gene variants implicated in CAS; *SETD1A* ^4^, *DDX3X* ^5^ and *SETBP1* ^5, 6^. We highlight chromatin organization and transcriptional regulation as critical biological mechanisms underpinning speech development.

The high confidence variants in this study were all located in genes previously associated with other common neurodevelopmental phenotypes including epilepsy, intellectual disability and ASD ^4, 5^. These complex speech and neurodevelopmental presentations match our current understanding of genes that have been associated with ASD, epilepsy and/or intellectual disability, where pleiotropy, or overlapping comorbid phenotypes, are common ^39^. However, for 15 of the 18 genes, this is the first time they have been specifically associated with CAS. Our work highlights the current bias in the literature to gene discovery cohorts across intellectual disability, autism and epilepsy relative to speech disorder. Hence, we have expanded the phenotypic spectrum for a number of genes previously implicated in neurodevelopmental disorders, linking them with a specific speech diagnoses, as well as markedly increasing the list of genes that should be prioritized for clinical testing in individuals with CAS.

Probands for whom we could provide a genetic diagnosis had a higher proportion of motor, language and cognitive impairments, secondary to the primary concern of CAS, compared to those probands without genetic diagnoses. We provide preliminary evidence for a threshold effect where monogenic conditions may be more likely when individuals with CAS have additional neurodevelopmental conditions, although further work on larger cohorts is needed to confirm this hypothesis. Only two probands (11%) with genetic diagnoses (*SETD1B* (ID10)*, TRIP12* (ID18)) had CAS without co-occurring neurodevelopmental disorder diagnoses. These findings expand the spectrum of phenotypes associated with these conditions. *SETD1B* has been previously associated with epilepsy, intellectual disability and language delay, and *TRIP12* has been associated with non-syndromic intellectual disability ^40^. Our data suggest that monogenic causes can underpin the less commonly occurring ‘isolated’ CAS phenotypes. We also reinforce the observation that, just as recent reports have suggested there are no ‘autism-(specific) genes,’^39^ it appears ‘speech-specific’ monogenic conditions are also rare. This has recently been acknowledged for individuals with *FOXP2* variants, where the phenotypic spectrum has been expanded from a relatively specific speech condition to include learning difficulties in at least some of the affected individuals ^41, 42^.

In terms of the biological pathways associated with speech disorder, we found a significant over- representation of perturbed chromatin and transcriptional regulation pathways, consistent with prior studies ^4, 5^. The five chromatin-related genes habouring high confidence variants in the current cohort were significantly co-expressed during brain development and are co-expressed with similar genes previously implicated in CAS ^4, 5^. *KDM5C* encodes a histone demethylase involved in regulation of gene expression ^43^ and DNA methylation ^44^, and LoF mutations in this gene have been shown to cause intellectual disability in females ^45^. *BRPF1,* encoding a histone acetyl transferase, has also been associated with intellectual disability and dysmorphic features ^46^. *SETD1A* encodes a histone methyltranferase and has previously been associated with schizophrenia, intellectual disability, and speech and/or language delays^33^. *SETD1B* is also a histone methyltransferase associated with neurodevelopmental disorder ^47^. *PHF21A* is a member of the *BRAF35*/histone deacetylase complex that mediates repression of neuron-specific genes ^48^ and has previously been associated with ASD and intellectual disability ^49^. *HNRNPK* encodes an RNA-binding protein known to interact with many molecular partners in multiple processes that regulate gene expression: chromatin remodeling, transcription, and mRNA splicing, translation, and stability ^50^. *De novo* truncating variants in *HNRNPK* have been shown to cause Au-Kline neurodevelopmental syndrome including intellectual disability, ADHD, speech impairment, cardiac anomalies and a variety of dysmorphic features ^37^. These results support earlier findings ^4, 5^ that chromatin modifiers and transcriptional regulators are critical for speech development. More generally, chromatin modifiers play important roles in neurodevelopmental disorders as they are key regulators in progenitor expansion, differentiation, cell-type specification, migration and maturation, with early errors in chromatin remodelling known to impact development of brain networks.^51^

Other genes confirmed to harbour pathogenic variants in CAS and associated with chromatin organization and transcriptional regulation were not as highly co-expressed during brain development. Among these, *DDX3X,* regulates gene splicing and is associated with neurodevelopmental disorders characterised by intellectual disability, ASD ^52^ and more recently, CAS^5^. *RBFOX3*, a gene showing neuron-specific expression, is also involved in splicing and has been associated with epilepsy aphasia syndrome and impaired language ^53^. *PURA* is implicated in the control of both DNA replication and transcription ^54^, and PURA syndrome is noted to include ‘absent speech’ as a feature of the condition ^55^. *ZBTB18* encodes a transcriptional repressor shown to play a critical role in orchestrating brain development, and has been associated with non-syndromic intellectual disability^56^.

The remaining candidate genes that we newly implicate in CAS (*TAOK2, SPAST* and *ARHGEF9*) encode proteins with distinct functions. *TAOK2* is located in the 16p11.2 deletion region, a well-recognised CNV associated with CAS, among other neurodevelopmental phenotypes ^57, 58^. The protein encoded by *TAOK2* has established roles in dendrite growth and synapse development ^59^. *ARHGEF9* encodes collybistin, a brain-specific guanine nucleotide exchange factor. The gene has also been implicated in X-linked epileptic encephalopathy and neurodevelopmental disorder, where there is skewed X-inactivation in favour of the abnormal X-chromosome ^60^. Finally, *SPAST* promotes microtubule growth in the cytoskeleton, playing an important role in neuronal development, and has been implicated in spastic paraplegia with dysarthric speech ^61^. Although previous reports describing individuals with *SPAST* variants have not thoroughly characterised speech in the early years of life, it is possible that CAS was part of the early profile of such cases. A pattern of CAS alongside dysarthria is not uncommon in other genetic forms of severe speech disorder, for example in Koolen de Vries Syndrome, ^62^ *SETBP1*- haploinsufficiency disorder ^6^, or *EBF3*-related core motor disturbance and ataxia.^5^

Whilst we were able to attain a diagnostic rate of 26%, other highly penetrant risk variants may have remained hidden due to our strict definition of high confidence variants, which was largely based on ACMG guidelines. By definition, for an identified variant to be deemed high confidence, a causative link to relevant disorder must be already established. Hence it is likely that a proportion of our of our low confidence candidate variants are truly causal but currently lack sufficient evidence. Identification of additional variants in these low confidence genes in future cohorts of individuals with CAS would elevate these findings to declare these genes as true CAS genes. Currently, they provide candidate genes for future studies. We also performed an expanded variant identification analysis including short tandem repeats which yielded no hits, suggesting at this time, that they do not play a major role in CAS ^63, 64^.

Our data confirm that a substantive proportion of children with CAS or related severe speech disorders may have a monogenic condition. As such CAS can be viewed as a critical clinical indicator for single gene disorders, due to its sensitivity as a rare phenotype (1 per 1000) ^4^, relative to more common speech diagnoses such as articulation or phonological disorder (1 in 20)^65^. While some individuals may have relatively ‘specific’ CAS in the absence of other neurodevelopmental disorders, our findings support the increasing overlap between genes conferring risk for a range of neurodevelopmental disorders including CAS, epilepsy, ASD and intellectual disability. This observation is important because well defined speech diagnoses are not typically reported in published clinical studies where the focus lies on other diagnoses like intellectual disability, epilepsy or ASD. If there is mention of speech or language impairment, the phrase ‘speech delay’ is typically used, which is a highly non-specific term that could imply general language understanding or expression difficulties (e.g. semantic or syntactic issues), and hence may not even be referring to ‘speech’ impairment itself (e.g. difficulty with producing speech sounds) ^12^. Thus, our work highlights the importance of specifically describing speech phenotypes in genotype-phenotype correlation studies and in considering a core diagnosis of CAS as a red flag for a monogenic condition. Understanding the aetiological basis of CAS is critical to end the diagnostic odyssey, identify comorbidities and ensure patients are poised for precision medicine trials.

## Supporting information

Supplementary methods and results

## Data Availability

All data produced in the present study are available upon reasonable request to the authors and pending Human Research Ethics Committee of the Royal Children's Hospital Melbourne, Australia, approval.

## Acknowledgments

The authors thank the families for participating in this study.

## Study funding

This work was funded by a National Health and Medical Research Council (NHMRC) Centre of Research Excellence Grant (CRE-SLANG; 1116976, AM, MH, IS, MB, DA, SEF) and an NHMRC project grant (1160893, AM, MB, DA, MH, SEF). AM was supported by an NHMRC Practitioner Fellowship (1105008) and Investigator grant (1195955). KB was supported by Australian Research Council Future Fellowship (120100355). MB was supported by a NHMRC Senior Research Fellowship (1102971) and a NHMRC Investigator grant (1195236). IS was supported by an NHMRC Investigator grant (1172897). SEF is supported by the Max Planck Society.

Additional funding was provided by the Independent Research Institute Infrastructure Support Scheme and the Victorian State Government Operational Infrastructure Program.

## Disclosure

The authors have no conflicts of interest to declare.

Supplementary information is available at Molecular Psychiatry’s website

## Notes

### Competing Interest Statement

The authors have declared no competing interest.

### Author Declarations

Ethics committee Human Research Ethics Committee of the Royal Children's Hospital Melbourne, Australia, gave ethical approval for this work.

