## Supplementary methods and results for "Genetic aetiologies for childhood speech disorder: novel pathways co-expressed during brain development"

### Supplementary Materials

### *Supplementary Methods*

#### Short tandem repeats

CAS probands were interrogated for the presence of expanded short tandem repeats (STRs) known to be associated with disease (Supplementary Table 3) using ExpansionHunter (v4.0.2) ^1^ and exSTRa (v. 1.1.0) ^2^ as well as a genome-wide search for novel, potentially disease-causing STR expansions using ExpansionHunter Denovo (EHdn v0.9.0) ^3^. For each locus, we reviewed plots of the STR size estimated by ExpansionHunter and the empirical cumulative distribution function (eCDF) generated by exSTRa, looking for outlier samples with one or both alleles, depending on the inheritance model, above the established disease-causing threshold for each locus (Supplementary Table 3). We performed outlier testing on the STR profiles generated by EHdn by designating all CAS probands and affected parents as “cases” and unaffected parents as “controls”. STR expansions with z>5 were filtered to remove expansions that lie in intergenic regions and remove regions with 500 base pairs of expansions with the same repeat motif identified by EHdn in the expanded 1000 Genomes cohort ^4^ (after lifting over coordinates from hg38 to hg19). Candidate STR expansions called by EHdn that remained after filtering were manually reviewed by performing a targeted follow up search using ExpansionHunter and exSTRa, as described above.

#### Calculation of polygenic risk scores

ASD, cleft palate, and word reading ability are three neurodevelopmental disorders with potential for phenotypic and genotypic overlap with CAS through shared genes such as *DDX3X*, *MEIS2,* *SETBP1* ^5, 6^ for ASD, or through potential phenocopy for cleft palate ^7^, and word reading having a phenotypic overlap with speech ^8^ . ASD, cleft palate and word reading also have polygenic risk components as part of their genetic aetiologies which have been defined in multiple, well-powered GWAS. Polygenic risk scores (PRS) summarise these risk profiles. PRS for Autism Spectrum Disorder (ASD), cleft palate and word reading were calculated by using the additive model with weighting by the effect sizes (betas) for the most highly associated risk alleles for each locus defined in the relevant publications describing genome-wide association study (GWAS) results for these three traits ^8-10^. Betas were extracted from Table 1 of Grove et. al., for ASD, table S3 of Howe et. al, for cleft palate and table S7 of Eising et. al. for word reading. All PRS were based on GWAS from European ancestry only analyses. Only genome-wide significant risk alleles (p<5e-08) were used to construct each PRS. If more than one genotype was missing in a sample, that sample was removed, otherwise the uncalled genotype was set to the reference genotype in a simple imputation procedure. PRS distributions were compared between CAS patients and matched controls. The trio sequencing design permitted the generation of pseudo-controls which were generated using the non-transmitted alleles from the parents, to give an ancestrally matched control population ^11^. The PRSs in cases was compared to the pseudo-controls using two-sided t-tests, assuming unequal variance. Estimation of the power required for this analysis to achieve statistical significance was calculated using the difference in the mean PRS scores between cases and pseudo-controls with the larger standard deviation of the two groups used as the standard deviation for the power calculations.

#### Estimation of mitochondrial abundance

WGS data have previously demonstrated utility as a biomarker for mitochondrial abundance ^12^. Mitochondrial DNA (mtDNA) abundance, or copy number (CN), was calculated for each sample by comparing the average WGS coverage in mtDNA and to that in chromosome 1 as previously described ^12^. To assess the impact of mtDNA abundance in CAS patients we performed robust linear regression analysis within each sequencing batch. We conducted outlier analysis within each batch using the z-score of the log transformed estimate of mtDNA CN, reporting the outlier samples (z-score > 2, z-score < -2). We intersected identified outliers with our genetic findings.

#### Co-expression analysis

Gene co-expression analyses were undertaken as previously described (6), using normalized brain expression values (reads per kilobase of exon model per million mapped reads [RPKM]) from the BrainSpan Developmental Transcriptome dataset (Gencode v10 summarized to genes). (28) Samples were restricted to include those from fetal and infancy periods only (8 weeks post-conception to 10 months after birth; data for included samples, Supplemental table 4), and all available brain regions were used. Following sample restriction, we removed genes with expression values missing from >50% of samples, expression values of 0 RKPM for ≥50% of samples, or variance of expression across all samples <0.5. After filtering, data from a total of 15,392 genes across 280 samples from 24 individuals remained.

A matrix of weighted correlations was generated, using the log2 transformed expression values and weights determined as $1/\sqrt{n}$ , where n is the number of samples contributed by the respective individual. Correlation heatmaps were generated using the corrplot R package (version 0.84, available at github.com/taiyun/corrplot), with genes ordered by hierarchical clustering, using the median linkage method. Co-expression gene networks were constructed using the qgraph R package (Epskamp et al. 2012), as follows: using the distribution of pairwise correlations of all 15,392 genes in the cleaned dataset, a threshold of |ρ| > 0.647 was determined, corresponding to the absolute correlation value that the 5% most highly correlated genes exceeded. Networks were then constructed with edges drawn between genes with pairwise correlations exceeding this threshold. For the purposes of downstream analysis, a cluster is determined by each member gene having at least four of these positive significant correlations with each other members of the cluster.

Finally, we determined whether these genes were significantly more highly co-expressed in the brain than would be expected for a random set of genes. We used a Monte Carlo sampling approach to approximate the distribution of the median |ρ| for all sets of genes, by randomly sampling 5,000 sets of x genes, the same number of genes as present in our high-confidence set, and calculating the median |ρ| for each random gene set. We derived an eCDF based on these medians, to which we compared the observed median |ρ| of our high-confidence candidates.

#### Using co-expression analyses to prioritize genes of uncertain significance

Given that we were able to identify significant co-expression of genes involved in speech disorder, we sought to leverage these gene expression patterns to identify further candidate genes for CAS as had been performed previously for other NDDs ^13, 14^. Firstly, we examined genes containing the low-confidence variants identified in the present study, and secondly, sets of candidate genes identified through literature searches, with so far limited direct evidence of involvement in CAS, compiled as follows: a) genes containing low-confidence variants identified here and in our previous cohorts ^15^ ; b) a set of 22 lists of genes, overlapping cytogenetic structural variants which had been previously associated with speech disorders or had speech disorder as a feature of the associated phenotypic spectrum (Supplementary table 8 and 9) and c) a set of 81 candidate speech genes derived from a literature search (Supplementary table 2) .

Gene prioritization was based on connectivity to the 34 CAS genes noted earlier, drawn from this study and past cohorts ^15, 16^. A connectivity score $(C)$ was generated for all genes in the candidate list, based on the pairwise correlations between that gene and the genes in the reference set. This score was calculated as the sum of the absolute correlations in the top 5% of pairwise correlations, defined as |ρ| > 0.647.

$$C=\sum\left| \rho\right|I$$

$$where I=\left\{ \begin{aligned} 1 if \left| \rho\right|>0.647 \\ 0 otherwise \end{aligned} \right.$$

We then determined which genes in the candidate list should be prioritized, based on evidence of co-expression with the reference set, as follows: 5000 sets of genes were randomly selected, with each set containing the same number of genes as in the candidate gene list. Within each random gene set, a connectivity score with the reference set was generated for each gene. A false discovery rate (FDR) for each candidate gene was then determined as the average proportion of genes in each of the 5000 random sets, that would be prioritized, assuming a threshold score equivalent to the candidate gene’s connectivity score.

For the $i^{th}$ candidate gene, with connectivity score $C_{i}$:

$$FDR_{i}=\frac{\sum_{j}^{5000} P_{j}}{5000}$$

$$where P_{j} is the proportion of genes in random set j, with a connectivity score \geq C_{i}$$

We performed the prioritization analysis for each candidate gene list separately, and for each list we selected candidate genes with an FDR < 0.1 for prioritization.

### *Supplementary Results*

#### Short Tandem Repeats

No pathogenic STR expansions were identified. We followed up two candidate STR expansions from a shortlist of 40 candidates that survived filtering and excluded them based on manual review.

#### Calculation of polygenic risk scores

PRS distributions were compared between the probands and the pseudo controls (generated from the untransmitted haplotype using the parents’ genotypes) (n=75) for ASD, cleft palate, and word reading traits ^8-10^. No significant differences (p<0.05) were observed in the PRS between the probands and pseudo controls (two sample t-test, assuming unequal variances) (Supplementary Fig. 2A-C). Both ASD and cleft palate demonstrated a trend for increased risk in CAS probands compared to controls with ASD close to nominal significance (p=0.054).

Using these data we conducted a power study. To achieve power of 0.8 at 0.05 significance the following minimum sample sizes will be required for ASD, cleft palate and word-reading respectively: 155, 405 and 206, demonstrating that we were underpowered to detect such differences with the current cohort size of 75.

#### Estimation of mitochondrial abundance

Mitochondrial dysfunction has been proposed to play a role in multiple nueurodevelopmental disorders (NDDs) ^17^. Mitochondrial copy number analysis applied to the combined cohort of WGS samples from ^15^ and the current cohort showed significantly increased (z-score > 2) in mtDNA CN in 6/81 probands, 2/29 affected relatives and 2/123 unaffected relatives. Also 2/123 unaffected relatives (parents) showed a significant decrease in copy number (z-score < 2). Of the six probands showing an increase in mtDNA CN, two have a variant associated with CAS. Family 2 (trio with unaffected parents) has a pathogenic variant in the *DDX3X* gene. The *DDX3X* gene has been previously associated with mitochondrial protein quality control ^18^. Family 8 (trio with unaffected parents) has a pathogenic variant in the *HNRNPK* gene. The *HNRNPK* gene has been previously associated with mitochondrial function through the correct splicing of Mitochondrial Ribosomal Protein L33 (*MRPL33*) ^19^. Hence mtDNA CN does not appear to be a biomarker for CAS.

#### Using co-expression analyses to prioritize genes of uncertain significance

Firstly we re-examined the set of low-confidence variants identified in the present study (Table 3b and c), and the low confidence variants identified in our previous cohort ^15^. Four genes in total were identified for prioritization (FDR<0.1, *BRWD3, MCMBP*, *TET3, and ZKSCAN1*). All are associated with chromatin organization and/or DNA binding, also captured by our empirical gene co-expression networks ^20-22^.

Next, we sought to use the new model to help us understand a set of cytogenetic structural variants which had been previously associated with speech disorders or had speech disorder as a feature of the associated phenotypic spectrum (Supplementary tables 8 and 9). We hypothesised that particular genes in each cytogenetic region of interest was likely to be the driver of the CAS part of the phenotype. We considered a set of genes defined by each structural variation derived from the literature. We considered 21 cytogenetic regions (some which overlap) with a range of 10 to 222 candidate genes and identified any significantly (FDR <0.1) prioritized candidates (Table 4) (Figure 4E). We identified more than one candidate for 18/21 regions. Some of the genes that we were able to prioritize already had strong evidence associating them with severe speech disorders and ID. For example, *ARID1B*, part of the 6q25.3 microdeletion, has been identified by ^23^ as likely causal, while *ERC1*, part of the 12p13.33-p13.32 interstitial deletion, has also been implicated as the key causal gene for the speech disorder component of the phenotype associated with this deletion ^24^.

In addition to identifying genes already implicated in CAS, prioritization analysis was able to provide evidence for other potential CAS candidates. *BCL9,* identified through prioritization analysis, was proposed as one of two likely causative candidates in the 1q21.1 microduplication syndrome ^25^. A deletion located at 7q31.2-q31.31, which includes *CTTNBP2* as well as 14 other genes, has previously been associated with a speech and language disorder ^26^. Our prioritization analysis identified *CTTNBP2*, also identified by the authors as the CAS gene in this deletion, despite its proximity to *FOXP2*, a well-known CAS gene.

Finally, we examined a set of 81 candidate speech genes derived from a literature search (Supplementary Table 2). Of these candidate genes, prioritization analysis highlighted *AUTS2*, *SETX*, and *CNTNAP1* at an FDR<0.1 threshold, as putative CAS genes based on gene co-expression. These genes were suggested as candidates in earlier small cohort family high-throughput sequencing studies ^27, 28^.

### Supplementary Figures

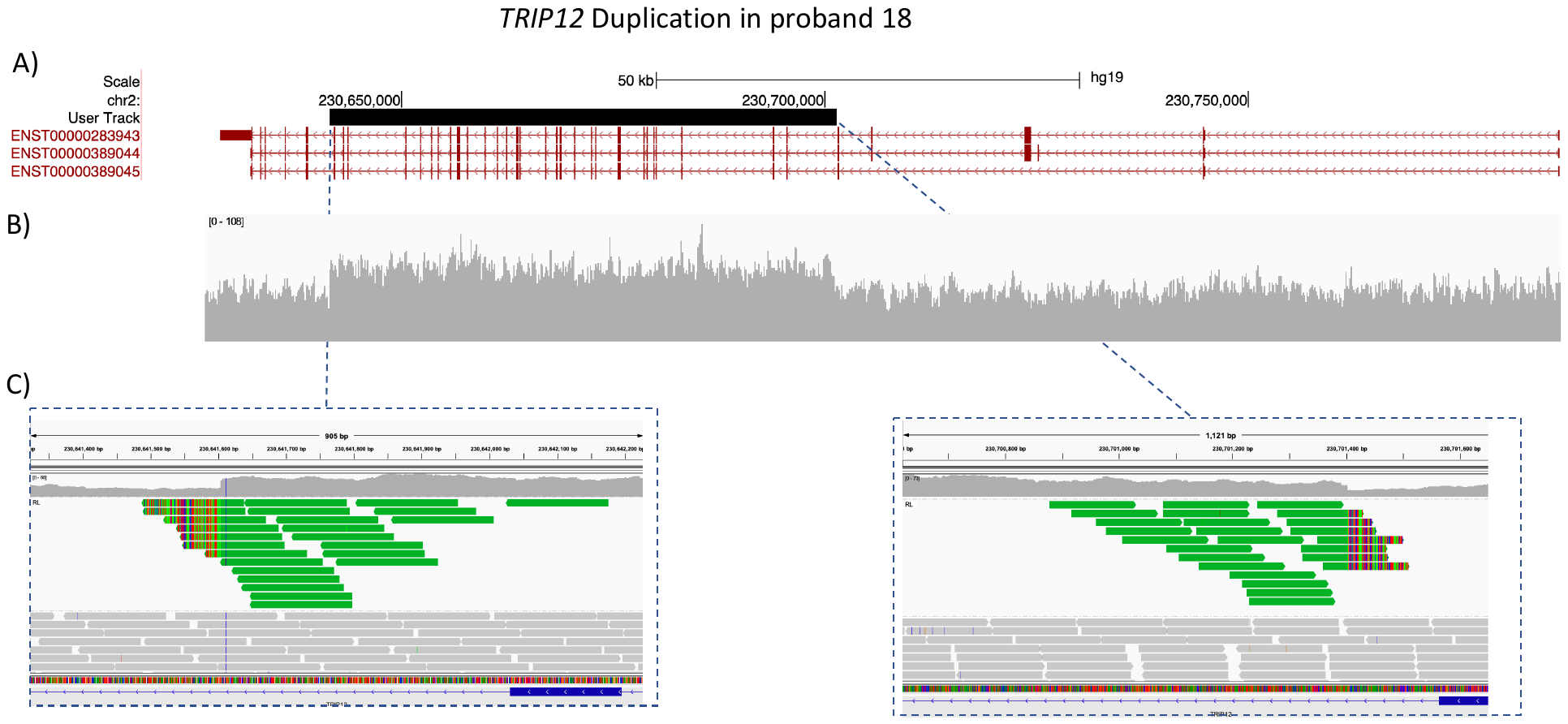

Supplementary Fig 1. Visualization of the multi-exonic duplication in the *TRIP12* gene (exons 7 to 37 of 42 in the MANE isoform NM_001348323.3; predicted out of frame) in proband 18. A) The location of the duplication in the hg19 build of the UCSC genome browser (chr2:230,641,603-230,701,402). The Manta predicted duplicated region (black). Three of the Ensembl defined transcripts for *TRIP12* (red). B) IVG visualization of base resolution read depth. C) IGV visualization of the ends of the tandem duplicated region. Green reads - mate pairs are aligned to the other end of the break. Green reads with colors strips indicate that part of the read is not aligned to the reference genome, rather aligned to the other end of the break (derived from individual read secondary alignment details).

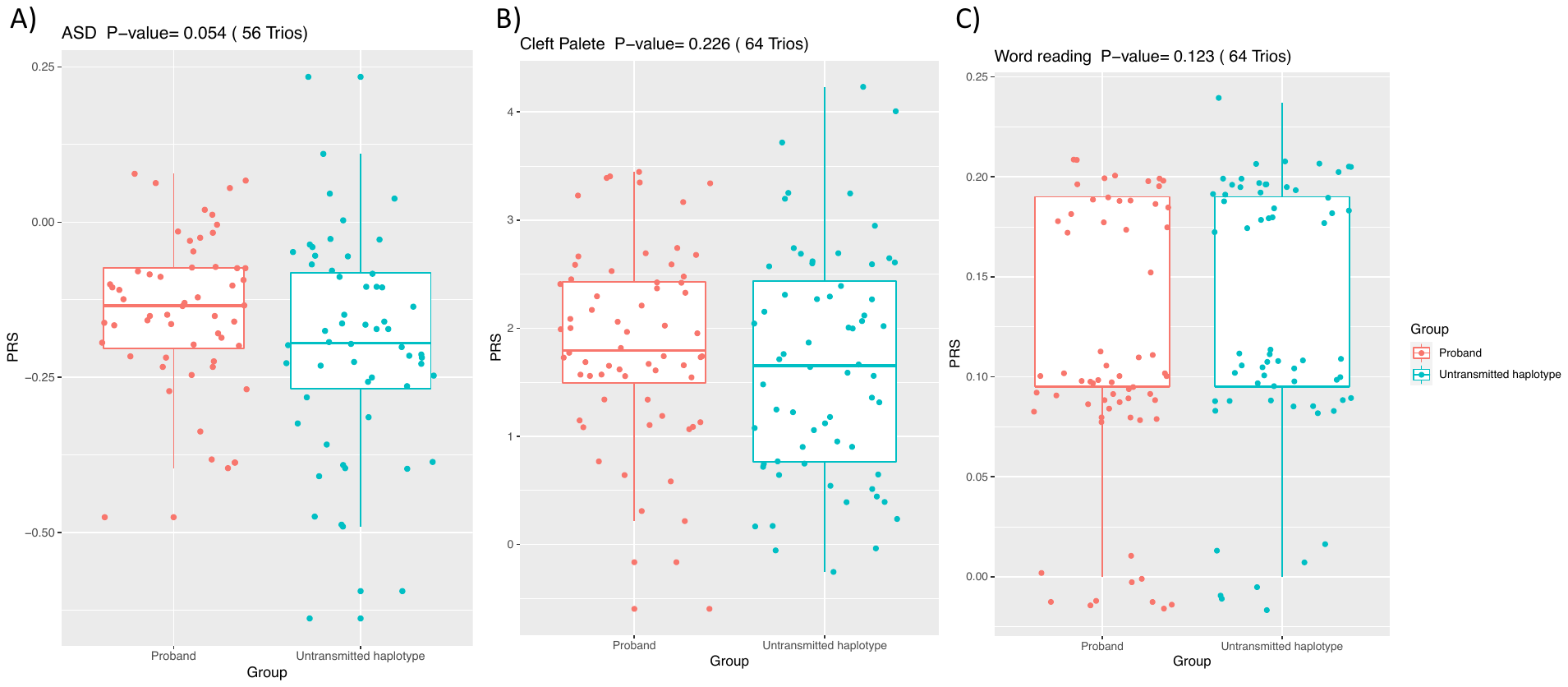

Supplementary Fig. 2. Distribution of Polygenic Risk Scores (PRS) for probands and pseudo-controls for A) Autism Spectrum disorder B) Cleft palate C) Word Reading. Boxplots represent interquartile range with the medium represented in bold. Whiskers represent the largest/smallest value within 1.5x the interquartile range outside the interquartile range.

### Supplementary Tables

Supplementary Tables 1-7 in Excel spreadsheet named KaspiSpeech_SupplementaryTables.xlsx.

Supplementary Table 8 - Large copy number regions

| Region | Gene List source* | Reference: |
| --- | --- | --- |
| 1q21.1 microdeletion | UCSC | ^25^ |
| 2p15 | org.Hs.eg.db | ^29^ |
| 2p16.1 | org.Hs.eg.db | ^30^ |
| 5q14.3 | org.Hs.eg.db | ^31^ |
| 5q14q21.1 | Paper | ^15^ |
| 6p21.3 | org.Hs.eg.db | ^32^ |
| 6q25.3 | org.Hs.eg.db | ^23^ |
| 7q11.23 | org.Hs.eg.db | ^33^ |
| 7q31.1 | org.Hs.eg.db | ^34^ |
| 7q31.2-q31.31 | Paper | ^26^ |
| 10q21.2-22.1 | UCSC | ^35^ |
| 12p13.33-p13.32 | UCSC | ^24^ |
| 15q13.3 microdeletion | UCSC | ^36^ |
| 15q14 | UCSC | ^37^ |
| 15q26.3 | org.Hs.eg.db | ^38^ |
| 16p11.2 | org.Hs.eg.db | ^39^ |
| 16p11.2 microdeletion | UCSC | ^40^ |
| 16p13.2 | org.Hs.eg.db | ^41^ |
| 16q23.2 | org.Hs.eg.db | ^42^ |
| 17p11.2 | org.Hs.eg.db | ^43^ |
| 19q13.11 | org.Hs.eg.db | ^44^ |

*Paper – gene names extracted directly from cited paper

*UCSC – gene names extracted from the USCS genome browser based on the coordinates and human genome build given in the paper

*org.Hs.eg.db – Bioconductor Genome wide annotation for Human library based on Entrez Annotation (<https://bioconductor.org/packages/release/data/annotation/html/org.Hs.eg.db.html>)
