## Supplemental Table 1 for "Genetic aetiologies for childhood speech disorder: novel pathways co-expressed during brain development"

Supplementary Table 1a - Medical and neurodevelopmental features of patients without confirmed variants

| **Family** | **Age, y;m^~^** | **Sex** | **Core speech phenotype** | **Gross motor delay** | **Fine motor delay** | **Vision impaired**^#^ | **Hearing loss**^#^ | **MRI findings**^#^ | **Seizures**^#^ | **Other NDD**^#^ | **Dysmorphic features**^#^ | **Other medical**^#^ |
| --- | --- | --- | --- | --- | --- | --- | --- | --- | --- | --- | --- | --- |
| **19** | 3-5 | M | CAS, phonological delay | Y | Y |  |  |  |  |  |  |  |
| **20** | 3-5 | F | CAS | NA | NA |  |  |  |  |  |  |  |
| **21** | 3-5 | M | Phonological disorder | Y | Y |  |  |  |  |  |  |  |
| **22** | 6-8 | M | CAS | N | N |  |  |  |  |  |  |  |
| **23** | 3-5 | M | CAS | Y | Y |  |  |  |  |  |  |  |
| **24** | 6-8 | M | CAS | Y | Y |  |  |  |  |  |  |  |
| **25** | 12-14 | F | CAS | Y | Y |  |  |  |  |  |  |  |
| **26** | 12-14 | M | CAS, Dysarthria | Y | Y |  |  |  |  |  |  |  |
| **27** | 6-8 | M | CAS | N | N |  |  |  |  |  |  |  |
| **28** | 3-5 | M | CAS, Phonological disorder | N | N |  |  |  |  |  |  |  |
| **29** | 6-8 | F | CAS | Y | Y |  |  |  |  |  |  |  |
| **30** | 3-5 | M | CAS | Y | Y |  |  |  |  |  |  |  |
| **31** | 3-5 | F | Dysarthria, phonological disorder, history of stuttering | Y | Y |  |  |  |  |  |  |  |
| **32** | 3-5 | F | CAS | N | Y |  |  |  |  |  |  |  |
| **33** | 6-8 | M | CAS | Y | Y |  |  |  |  |  |  |  |
| **34** | 6-8 | F | CAS | Y | Y |  |  |  |  |  |  |  |
| **35** | 9-11 | M | CAS | Y | N |  |  |  |  |  |  |  |
| **36** | 3-5 | M | CAS | N | N |  |  |  |  |  |  |  |
| **37** | 3-5 | M | CAS | Y | Y |  |  |  |  |  |  |  |
| **38** | 3-5 | M | CAS | N | N |  |  |  |  |  |  |  |
| **39** | 6-8 | M | CAS | N | N |  |  |  |  |  |  |  |
| **40** | 6-8 | M | CAS | N | Y |  |  |  |  |  |  |  |
| **41** | 3-5 | M | CAS | N | Y |  |  |  |  |  |  |  |
| **42** | 6-8 | M | CAS | Y | Y |  |  |  |  |  |  |  |
| **43** | 3-5 | M | Phonological disorder | Y | N |  |  |  |  |  |  |  |
| **44** | 3-5 | F | CAS | N | N |  |  |  |  |  |  |  |
| **45** | 3-5 | M | CAS | N | Y |  |  |  |  |  |  |  |
| **46** | 3-5 | M | CAS | N | Y |  |  |  |  |  |  |  |
| **47** | 3-5 | M | CAS, Phonological delay | N | N |  |  |  |  |  |  |  |
| **48** | 3-5 | M | CAS | N | N |  |  |  |  |  |  |  |
| **49** | 3-5 | M | CAS | N | Y |  |  |  |  |  |  |  |
| **50** | 3-5 | M | CAS | Y | Y |  |  |  |  |  |  |  |
| **51** | 3-5 | M | Features of CAS | N | N |  |  |  |  |  |  |  |
| **52** | 9-11 | M | CAS | Y | Y |  |  |  |  |  |  |  |
| **53** | 6-8 | M | CAS | Y | Y |  |  |  |  |  |  |  |
| **54** | 6-8 | M | Phonological delay, features of CAS & dysarthria | Y | Y |  |  |  |  |  |  |  |
| **55** | 3-5 | M | CAS, phonological disorder | Y | Y |  |  |  |  |  |  |  |
| **56** | 3-5 | M | CAS, phonological disorder | N | Y |  |  |  |  |  |  |  |
| **57** | 6-8 | M | CAS | N | N |  |  |  |  |  |  |  |
| **58** | 3-5 | M | CAS | N | N |  |  |  |  |  |  |  |
| **59** | 6-8 | F | CAS | Y | Y |  |  |  |  |  |  |  |
| **60** | 3-5 | M | CAS, phonological delay | Y | Y |  |  |  |  |  |  |  |
| **61** | 3-5 | F | Features of CAS, phonological disorder | N | N |  |  |  |  |  |  |  |
| **62** | 6-8 | M | CAS | N | Y |  |  |  |  |  |  |  |
| **63** | 3-5 | M | CAS, phonological disorder | Y | Y |  |  |  |  |  |  |  |
| **64** | 3-5 | M | CAS | Y | Y |  |  |  |  |  |  |  |
| **65** | 6-8 | F | Dysarthria, features of CAS | Y | Y |  |  |  |  |  |  |  |
| **66** | 3-5 | M | Dysarthria, phonological disorder | Y | Y |  |  |  |  |  |  |  |
| **67** | 3-5 | M | Phonological disorder, features of CAS | Y | Y |  |  |  |  |  |  |  |
| **68** | 3-5 | M | CAS | Y | Y |  |  |  |  |  |  |  |
| **69** | 6-8 | F | CAS | N | N |  |  |  |  |  |  |  |
| **70** | 3-5 | F | CAS | Y | Y |  |  |  |  |  |  |  |

ASD, Autism spectrum disorder; CAS, Childhood apraxia of speech; DCD, Developmental coordination disorder; F, Female; GDD, Global developmental delay; ID, Intellectual disability; M, Male; N, Feature not present; NA, Not assessed; NDD, neurodevelopmental disorder; Y, Feature present; *Patient has a history of conductive hearing loss, treated with grommets, hearing was normal at the time of assessment, ^#^These details have been removed to avoid the identification of patients, **^~^**Ages have been denoted in 2-year bands to avoid the identification of participants.

Supplementary Table 1b: Extended linguistic phenotype and educational setting of patients without confirmed variants

| **Family** | Oral motor impairment | History of feeding issues | Language: receptive* | Language: expressive* | Reading deficits | Spelling deficits | Speech pathology | IQ^#^ | Education setting |
| --- | --- | --- | --- | --- | --- | --- | --- | --- | --- |
| **19** | Y | Y | Average | Mild | N | Y | Y |  | Mainstream |
| **20** | Y | NA | Average | Mild | NA | NA | Y |  | Mainstream |
| **21** | N | Y | Average | Average | TY | TY | Y |  | Not yet at school |
| **22** | Y | N | NA | NA | NA | NA | Y |  | Mainstream |
| **23** | Y | Y | Y~ | Y~ | TY | TY | Y |  | Not yet at school |
| **24** | Y | N | Severe | Severe | Y | Y | Y |  | Mainstream |
| **25** | Y | N | Y~ | Y~ | Y | Y | Y |  | Mixed mainstream and specialist |
| **26** | Y | Y | Severe | Moderate | Y | Y | Y |  | Specialist |
| **27** | N | N | Average | Mild | N | N | Y |  | Mainstream |
| **28** | Y | N | Average | Average | TY | TY | Y |  | Not yet at school |
| **29** | Y | Y | Severe | NA | Y | Y | Y |  | Mainstream |
| **30** | N | Y | N~ | Y~ | TY | TY | Y |  | Not yet at school |
| **31** | Y | Y | Average | NA | TY | TY | Y |  | Not yet at school |
| **32** | Y | N | N~ | Y~ | Y | Y | Y |  | Mainstream |
| **33** | Y | N | Severe | Severe | Y | Y | Y |  | Mainstream |
| **34** | Y | N | Average | NA | Y | Y | Y |  | Mainstream |
| **35** | Y | N | Mild | Y~ | Y | Y | Y |  | Mainstream |
| **36** | Y | N | NS | NS | NA | NA | Y |  | Specialist |
| **37** | Y | N | NA | NA | TY | TY | Y |  | Not yet at school |
| **38** | Y | N | Average | Above average | TY | TY | Y |  | Mainstream |
| **39** | Y | N | Mild | Mild | NA | NA | Y |  | Mainstream |
| **40** | Y | N | Average | NA | NA | NA | Y |  | Mainstream |
| **41** | N | Y | Average | Mild | NA | NA | Y |  | Not yet at school |
| **42** | Y | Y | Mild | NA | Y | Y | Y |  | Mainstream |
| **43** | Y | N | Mild | NS | TY | TY | Y |  | Mainstream |
| **44** | Y | N | Average | Severe | TY | TY | Y |  | Mainstream |
| **45** | Y | N | Average | NA | TY | TY | Y |  | Not yet at school |
| **46** | Y | N | Above average | Average | TY | TY | Y |  | Mainstream |
| **47** | Y | Y | Above average | NA | TY | TY | Y |  | Mainstream |
| **48** | Y | N | NA | NA | TY | TY | Y |  | Mainstream |
| **49** | Y | Y | Average | Average | TY | TY | Y |  | Mainstream |
| **50** | Y | Y | Severe | NS | TY | TY | Y |  | Mainstream |
| **51** | Y | Y | Average | NS | TY | TY | Y |  | Mainstream |
| **52** | Y | N | Average | Average | N | N | Y |  | Mainstream |
| **53** | Y | Y | Moderate | Mild | Y | Y | Y |  | Mainstream |
| **54** | Y | Y | Average | Severe | Y | Y | Y |  | Mainstream |
| **55** | Y | N | Average | Average | TY | TY | Y |  | Mainstream |
| **56** | Y | Y | Above average | Average | TY | TY | Y |  | Mainstream |
| **57** | N | N | Average | NA | N | N | Y |  | Mainstream |
| **58** | Y | N | Average | Severe | N | N | Y |  | Mainstream |
| **59** | Y | Y | Average | Average | N | N | Y |  | Mainstream |
| **60** | N | Y | Severe | Severe | N | N | Y |  | Mainstream |
| **61** | N | N | NA | NA | TY | TY | Y |  | Mainstream |
| **62** | Y | NA | Severe | Severe | NA | NA | Y |  | NA |
| **63** | Y | N | Severe | Severe | TY | TY | Y |  | Mainstream |
| **64** | Y | N | N~ | NS | TY | TY | Y |  | Mainstream |
| **65** | Y | N | Severe | Severe | Y | Y | Y |  | Mainstream |
| **66** | NS | Y | Average | Average | TY | TY | Y |  | Mainstream |
| **67** | Y | N | NA | NA | TY | TY | Y |  | Specialist |
| **68** | Y | Y | Severe | Severe | TY | TY | Y |  | Not yet at school |
| **69** | Y | N | Average | Severe | N | N | Y |  | Mainstream |
| **70** | Y | NA | Severe | Severe | TY | TY | Y |  | Not yet at school |

FSIQ, Full scale IQ; N, no; NA, Not assessed; NS, unable to score assessment, abilities likely severe range; N~, based on subtest scaled scores; VCI: Verbal Comprehension Index; VSI: Visual Spatial Comprehension; PRI, Perceptual reasoning index; TY; not applicable, too young (< 5 years old) to assess literacy; Y, feature present.

*Language severity rated according to CELF-5 as follows: 86-114 average, 78-85 mild, 71-77 moderate, <70 severe.

^ Assessment not indicated by the family or treating physician; % Participant received ID diagnosis after recruitment.

#These details have been removed to avoid the identification of patients.
